# Reassessing COVID-19’s First Year: Estimating Infections and Tracking Pandemic Trends with Probabilistic Bias Analysis

**DOI:** 10.1101/2025.11.21.25338950

**Authors:** Harsh Vivek Harkare

## Abstract

**Background:** The novel SARS-CoV-2 virus, first identified in China in December 2019, rapidly spread worldwide, resulting in 114 million confirmed cases and 2.5 million reported deaths by February 28, 2021. Yet considerable uncertainty persists regarding the true scale of infections, as limited testing capacity and inconsistent surveillance substantially hindered accurate case detection. Robust estimates of infection burden are essential for understanding the pandemic’s full impact and guiding effective public health responses.

**Methods:** This study aimed to quantify the health burden of COVID-19 in India, Mexico, the United Kingdom, and the United States. A probabilistic bias analysis model is used to estimate the true number of SARS-CoV-2 infections, accounting for factors such as repeated testing, infection waves, and viral mutations to provide a more accurate assessment of the pandemic’s true scale in each country.

**Results:** By February 28, 2021, the estimated total COVID-19 infections across India, Mexico, the United Kingdom, and the United States reached 286.7 million - more than six times the reported cases. India had the highest estimated infections (129.3 million), followed by the United States (98.6 million), Mexico (44.9 million), and the United Kingdom (14 million). Detection rates varied significantly, with Mexico underreporting infections by a factor of 22, while the United Kingdom and the United States had the highest detection rates. Testing capacity played a key role, with high-income countries conducting over four times more tests per 1,000 people than lower-income nations.

**Conclusion:** This study demonstrating substantial underreporting of SARS-CoV-2 during the pre-vaccination period. These retrospective estimates provide a more accurate historical baseline for interpreting pandemic dynamics and remain valuable for assessing long-term health impacts and improving preparedness for future epidemics.

## INTRODUCTION

In December 2019, cases of pneumonia of unknown origin were reported in Wuhan, China. The virus responsible, later named severe acute respiratory syndrome coronavirus 2 (SARS-CoV-2), caused the disease known as COVID-19. Declared a pandemic by the World Health Organization (WHO) in March 2020, COVID-19 spread rapidly across the world, resulting in millions of infections and deaths. Despite widespread public health measures, the virus continued to spread due to missed infections, testing limitations, emerging variants, and inconsistent compliance with containment measures. Understanding the true extent of infections remains essential for effective public health responses.

Testing is a crucial tool for controlling the pandemic, enabling the identification and isolation of infectious individuals. Various testing methods, including RT-PCR, antigen, and seroprevalence tests, have been deployed globally. Mass testing has been proposed as a strategy to curb transmission while minimising societal disruptions (Li et al., 2020; Sheikh et al., 2020; Weissleder et al., 2020). Countries that implemented large-scale testing, such as Slovakia, demonstrated significant reductions in infection prevalence (Pavelka et al., 2021). However, challenges such as resource constraints, access barriers, and testing accuracy have hindered widespread adoption.

In the early months of the pandemic, testing was primarily limited to symptomatic individuals or those with travel histories. As the understanding of asymptomatic transmission grew, testing strategies evolved, with some countries adopting broader or universal testing approaches. Asymptomatic and mild infections, which often go undetected, contribute significantly to community spread, highlighting the need for mass testing (Rothe et al., 2020; Shaman et al., 2020). Despite improvements in testing capacity, many countries still lack the resources to implement frequent and widespread testing.

Given the limitations of direct testing, alternative approaches are needed to estimate true infection numbers. Mathematical models can help provide a more accurate picture of infections, aiding policymakers in making informed decisions. These models, when used alongside testing data, can improve estimates of infection rates, transmission dynamics, and disease severity, offering valuable insights for public health planning (Lourenço et al., 2020; Pourmalek et al., 2021).

This research aims to track the progression of COVID-19 over one year and estimate the true number of infections in India, Mexico, the United Kingdom, and the United States using a probabilistic bias analysis approach until February 28, 2021. This period marks a critical phase of the pandemic before the widespread emergence of viral mutations and the global rollout of vaccines, allowing for infection estimates without the confounding effects of evolving immunity and vaccination strategies. As the model is not designed to account for vaccine-induced immunity, focusing on this timeframe ensures that estimates reflect the natural progression of infections under early pandemic conditions.

## METHODS

This study estimates the cumulative number of SARS-CoV-2 infections in India, Mexico, the United Kingdom, and the United States using a probabilistic bias analysis method. The study period spans from March 1, 2020, to February 28, 2021, tracking the progression of the pandemic over one year. Infection trends were analysed by observing changes in detection rates and population infection rates, along with correlations between testing rates and detection rates.

The approach builds upon a reference study by Wu et al. (2020), which estimated SARS-CoV-2 infections in the United States over a 51-day period. The reference model used Monte Carlo simulations to correct for biases due to incomplete testing and test inaccuracy. Prior distributions were defined for key testing probabilities, including sensitivity, specificity, and test positivity rates. This study extends the methodology by incorporating monthly variations in test positivity rates, the impact of infection waves, and the emergence of variants. Additionally, adjustments were made to account for the possibility of individuals being tested multiple times over a longer estimation period.

Probabilistic bias analysis was selected as it models uncertainty more effectively than simple bias corrections, which assume fixed bias parameters. Instead, this approach defines bias parameters as random variables within specified probability distributions, ensuring that uncertainty in testing probabilities is adequately captured. A beta distribution was used for prior definitions due to its suitability for proportions such as sensitivity, specificity, and test positivity rates.

Prior distributions were informed by country- and time-specific literature, including studies on testing probabilities, SARS-CoV-2 clinical characteristics, and diagnostic test accuracy. Moderate-to-severe symptomatic cases were classified based on CDC guidelines, while mild-to-asymptomatic cases were separately modelled (CDC, 2021). Given the evolving nature of the pandemic, priors for later periods were informed by earlier studies where necessary. The impact of viral variants was incorporated by adjusting priors based on documented increases in transmissibility and infectivity.

Monte Carlo simulations were performed by randomly sampling from each prior distribution 10,000 times per month. Monthly test positivity rates were calculated as the cumulative number of new confirmed cases divided by the cumulative number of tests conducted in each month.

To estimate the total number of SARS-CoV-2 infections, the number of susceptible individuals rather than untested individuals was used as a key parameter. This modification accounts for repeated testing of individuals and avoids artificial suppression of estimated infections over time. Monthly estimates were computed iteratively, adjusting the number of susceptible individuals by subtracting estimated and confirmed infections from the total population.

Key assumptions included: (1) all tests were assumed to be conducted via RT-PCR using nasopharyngeal swabs, (2) testing occurred within two weeks of infection, (3) RT-PCR test specificity remained unaffected by variants, and (4) prior infection conferred lasting immunity, preventing reinfection. The final estimates include both confirmed cases and undiagnosed infections, with results reported as medians along with 2.5th and 97.5th percentiles.

Bias corrections were applied to account for incomplete testing and test inaccuracies. Adjustments were made to ensure that the estimated number of missed infections was not artificially inflated or suppressed. The estimation process was repeated 10,000 times, and the final estimates reflect a range of possible true infection counts. Given the increasing prevalence of SARS-CoV-2 over time, the share of missed infections was primarily attributed to insufficient testing rather than imperfect test accuracy in the early stages of the pandemic.

To explore the relationship between detection and testing rates, a correlation analysis was conducted using Pearson’s correlation coefficient. Monthly detection rates were compared with key testing indicators such as average daily tests per 1,000 people and test positivity rates. The analysis was performed separately for each country to account for differences in distribution and testing strategies.

### Data

COVID-19 testing and case data for all countries were sourced from the *Our World in Data* repository, compiled by the University of Oxford, which integrates information from institutions such as Johns Hopkins University, the European Centre for Disease Prevention and Control, and national governments (OWID, 2021). The dataset includes testing figures, hospitalisations, ICU admissions, test positivity rates, and socio-economic indicators such as population density, income levels, and healthcare capacity. Testing units varied by country, with India, the UK, and the US reporting the number of tests performed, while Mexico recorded the number of individuals tested, though multiple tests for the same individual were counted separately. Data sources included the Indian Council of Medical Research, the Mexican government, Public Health England, and the US Department of Health and Human Services, covering PCR and antigen tests, with minor contributions from serology and non-PCR methods.

## RESULTS

There exists a significant variation in the evolution of COVID-19 testing rates among the studied sample of countries (Figure 1). The United Kingdom and the United States undergo a steady increase in both the number of average daily tests administered per 1,000 of their population as well as in the total cumulative number of tests administered. By March 2021, both countries had administered enough tests to test their entire populations once over. On the other hand, India and Mexico only see a very small, stagnating rise in their testing capabilities. At its peak, the United Kingdom had a daily testing rate of 8.7 per 1,000 in February 2021. During the same time, Mexico had a daily testing rate of 0.19 per 1,000.

**Figure 1:**
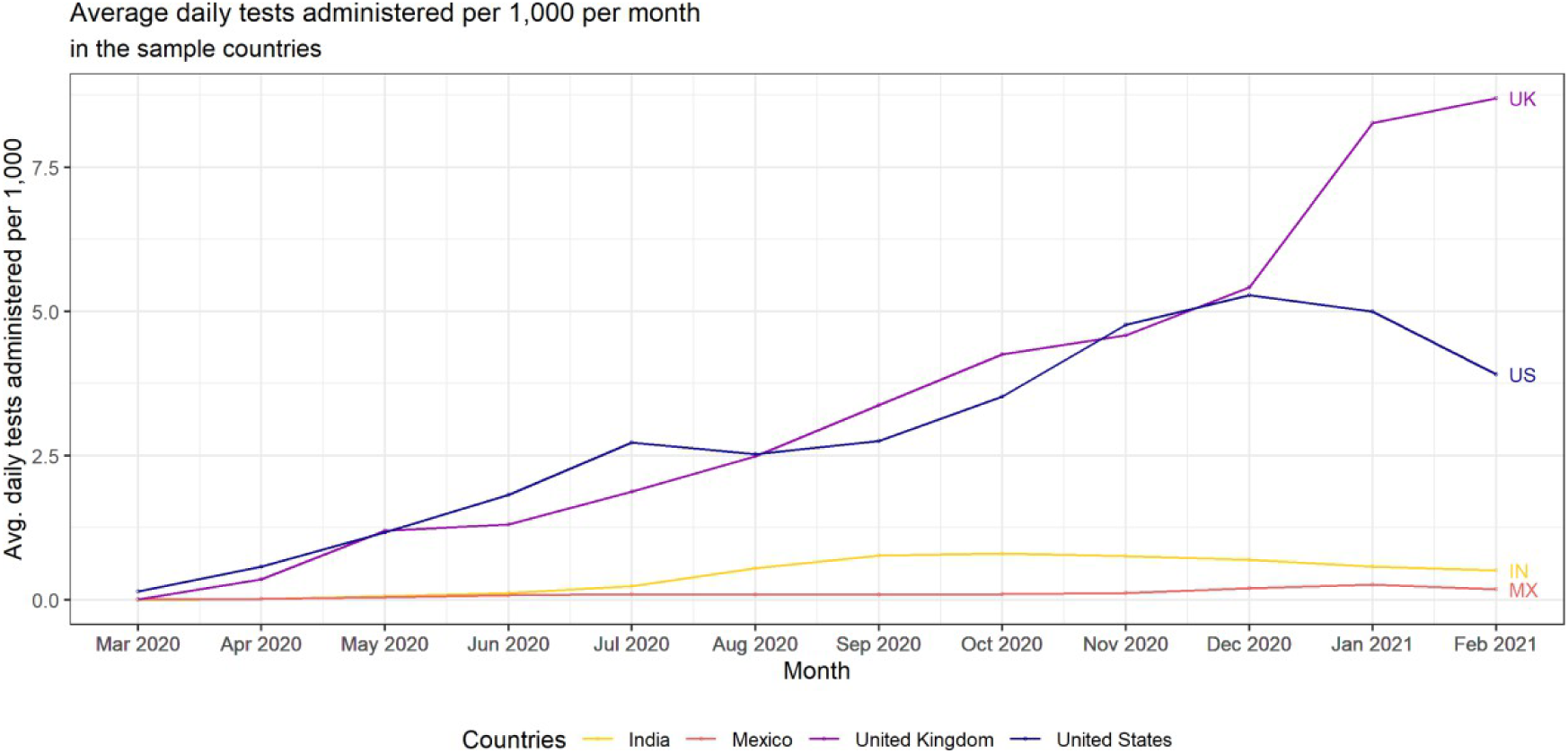
Average daily tests administered per 1,000 in the sample countries by month.

This trend in the rapid evolution of testing rates in richer countries can be seen on a wider scale as well. Looking at 15 of the top 20 countries with the highest reported COVID-19 cases as of February 28, 2021, we see a similar pattern (Figure S1). The average number of SARS-CoV-2 tests administered daily per 1,000 averaged over periods of one month is more than four times higher in countries with incomes above the average income per capita than those below it.

Corrected estimates of cumulative SARS-CoV-2 infections in India, Mexico, the United Kingdom, and the United States are presented along with the simulation interval in Figure 2. A summary of the estimated case counts, the population infection rates, and the detection rates is presented in Table 1. Simulation intervals of cumulative estimated infections, the population infection rate, and detection rate as well as monthly counts of estimated infections are provided as supplementary information (Figure S3, Table S5).

**Figure 2:**
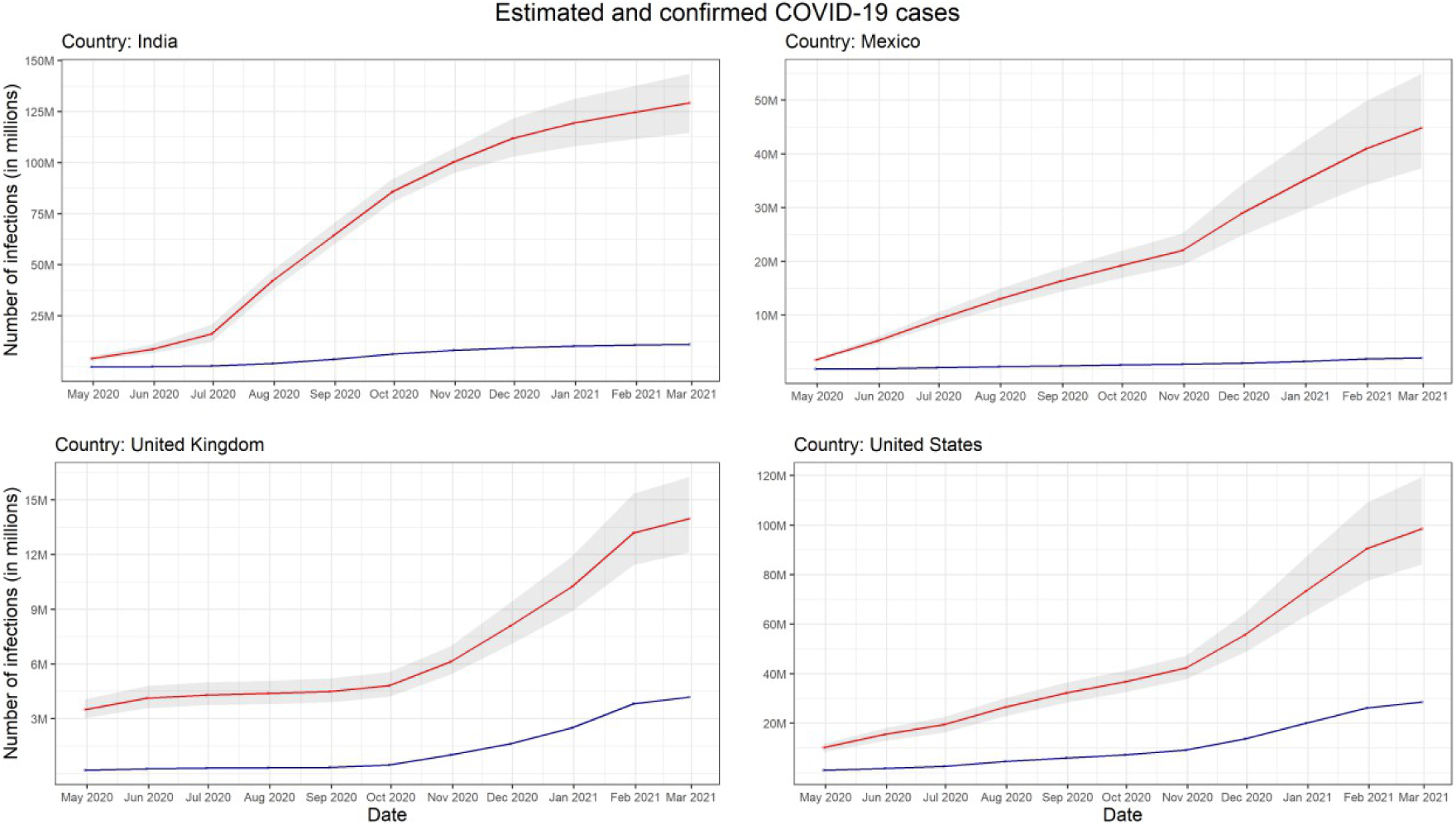
Cumulative estimated and confirmed cases in the sample countries. *Note: The red line represents the mean value of the estimated cases of SARS-CoV-2, the grey area represents the 95% confidence interval of the estimated cases, and the blue line represents the confirmed cases*.

**Table 1.**
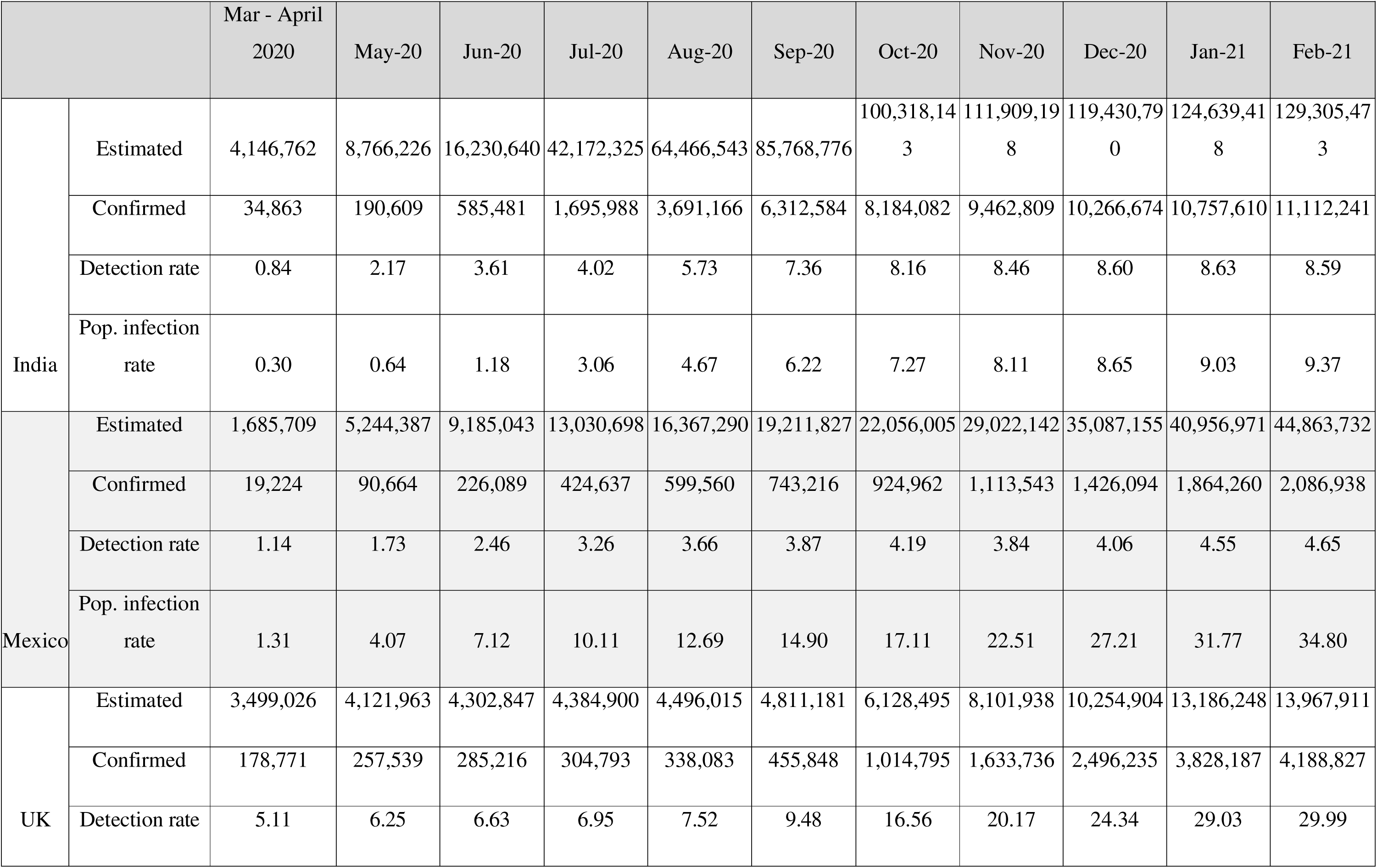
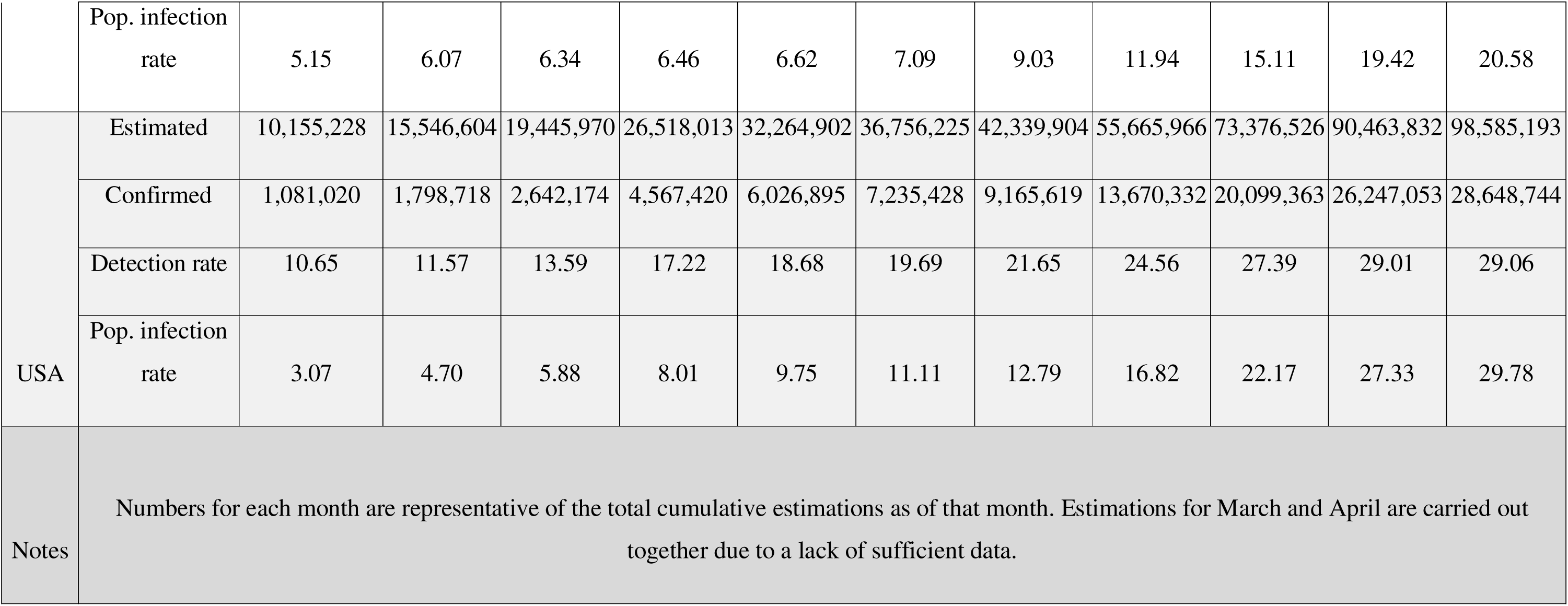
Monthly cumulative estimated infections, confirmed cases, detection rates, and population infection rates for COVID-19 in different countries

Between April 30, 2020, and February 28, 2021, the combined number of estimated cases in the sample countries increased from 19,486,726 to 286,722,309 - a factor of 15. Looking at specific countries, the estimated number of total infections by February 28, 2021 were highest in India at 129,305,473 infections. This is followed by the United States with 98,585,193 cumulative infections, Mexico with 44,863,732 infections, and the United Kingdom with 13,967,911 estimated infections. The larger simulation interval around the estimated SARS-CoV-2 infections for each country during the third wave of infections captures the greater uncertainty posed by the viral mutations.

For India, the estimated true number of infections were 11 times larger than the confirmed cases. According to the 95% simulation interval of infection estimates (114,617,975 - 143,626,761), the true number of infections were 10 - 13 times the confirmed cases. With 25 million infections, India is estimated to have reached a peak in infections in July 2020. Cumulative infections in Mexico were estimated to be 22 times higher than the reported numbers. Considering the 95% simulation interval (37,330,051 - 54,944,039), this number ranges between 18 - 26 times. Close to 7 million Mexicans are estimated to have been infected in November 2020. The estimated infections were 3.3 times higher than the confirmed cases in the United Kingdom. The 95% simulation interval for the number of estimated infections was 12,112,794 - 16,264,844. This would imply that the true number of infections were 2.9 - 3.9 times higher than the confirmed infections. The month of January 2021, with almost 3 million estimated infections, marked the peak of infections in the United Kingdom. Similar to the United Kingdom, the estimated infections were almost 3.5 times higher than the confirmed cases in the United States. 95% simulation interval of estimated infections for the United States was 84,052,809 - 119,505,262, corresponding to the estimated infections being 2.9 - 4.2 times higher than the reported infections. Of the overall total infections, 17.7 million were estimated to be infected in December 2020. Figure 3 presents the estimated infections per 1,000 as well as the size of missed infections in each of the studied countries.

**Figure 3:**
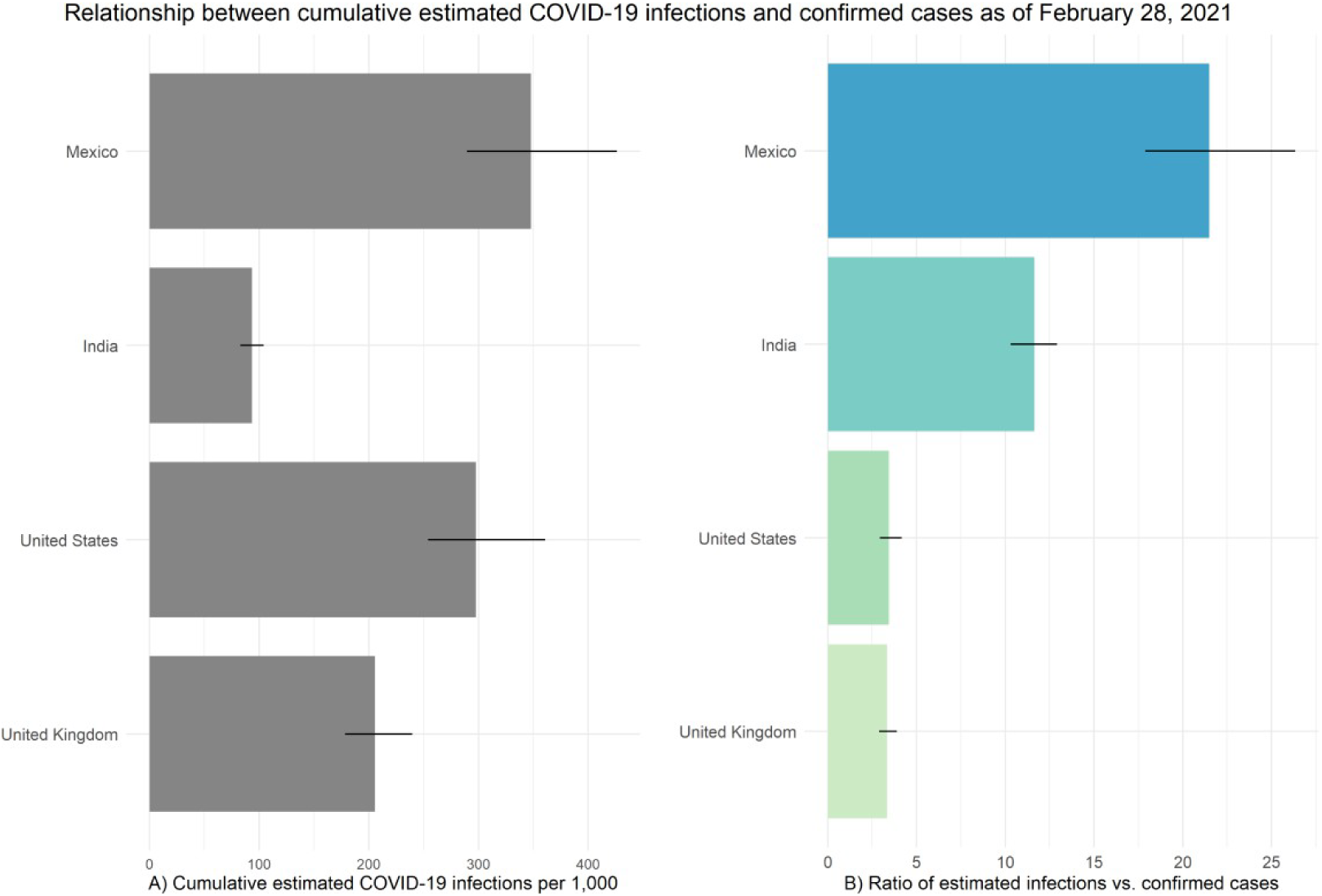
Estimated infections per 1,000 and size of missed infections

The United Kingdom and the United States have seen significant increases in their detection rates of true infections over time, even as they started with higher levels (Figure 4). By February 28, 2021, the detection rates for both countries were close to 30%, which translates to a 6-fold and 3-fold rise in detection rates over the studied period of one year for the United Kingdom and the United States respectively. The biggest jump in the detection rate for the United Kingdom can be seen for the month of October 2020, which was a time of high testing and low estimated infections. India also witnessed a considerable increase in detection rates for the first few months of the pandemic, but since October 2020 this growth has plateaued, similar to the pattern observed in the country’s testing rate. Over the one-year span of the study period however, the country witnessed the largest increase in detection rates among the studied set of countries. India’s detection rates increased from 0.84 in April 2020 to 8.6 in February 2021 - a factor of 10.2. Lastly for Mexico, even as the country underwent modest increases in its testing capacities, its detection rate increased four-folds over the study period; in part due to its low starting levels of 1.14.

**Figure 4:**
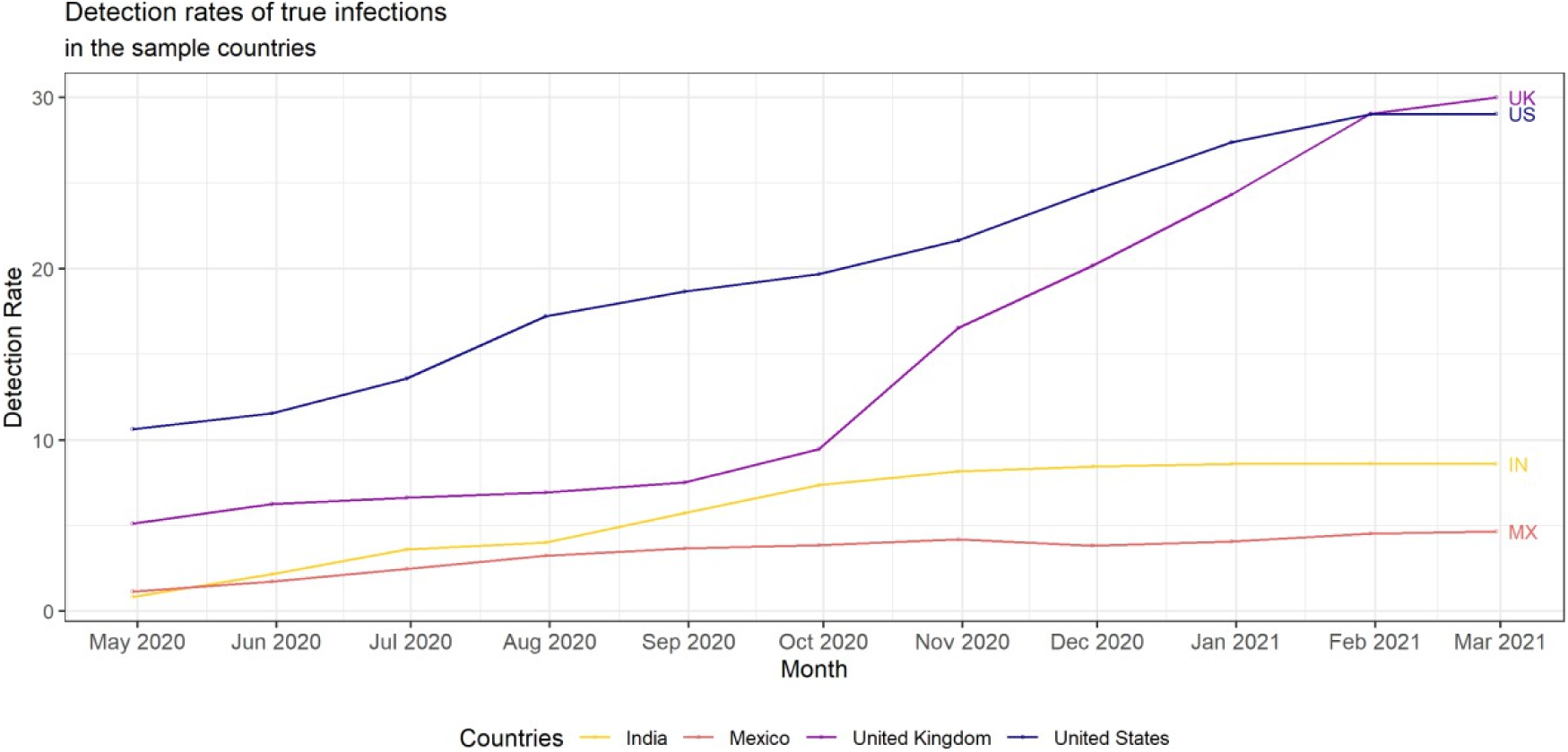
Trends in detection rates over time.

The largest increase in the share of population infected was documented for India, whose population infection rates increased by a factor of 31 from 0.3% in April 2020 to 9.4% in February 2021 (Figure S2). At the same time, India had the lowest share of total population infected. This was followed by Mexico with a 26-fold increase in the share of population infected. With 30% of its population infected, the United States saw only a 10-fold increase in its share of population infected. Finally, the rise in the population infection rate was the least for the United Kingdom. The share of infected individuals rose by a factor of 4 from 5.2% to 20.6%.

When plotting testing rates against monthly detection rates of the four sample countries, I observe statistically significant correlations for all countries (Figure S5). Looking at the relationship between the detection rate and the test positivity rate (Figure 5), I observe a statistically significant negative correlation for the United Kingdom (*r*(9) = -0.62, *p*< 0.05) and the United States (*r*(9) = -0.66, *p* < 0.05). India observes an insignificant positive correlation (*r*(9) = 0.42, *p* = 0.20), while Mexico observes a significant positive correlation (*r*(9) = 0.76, *p* < 0.05) between the two variables.

**Figure 5:**
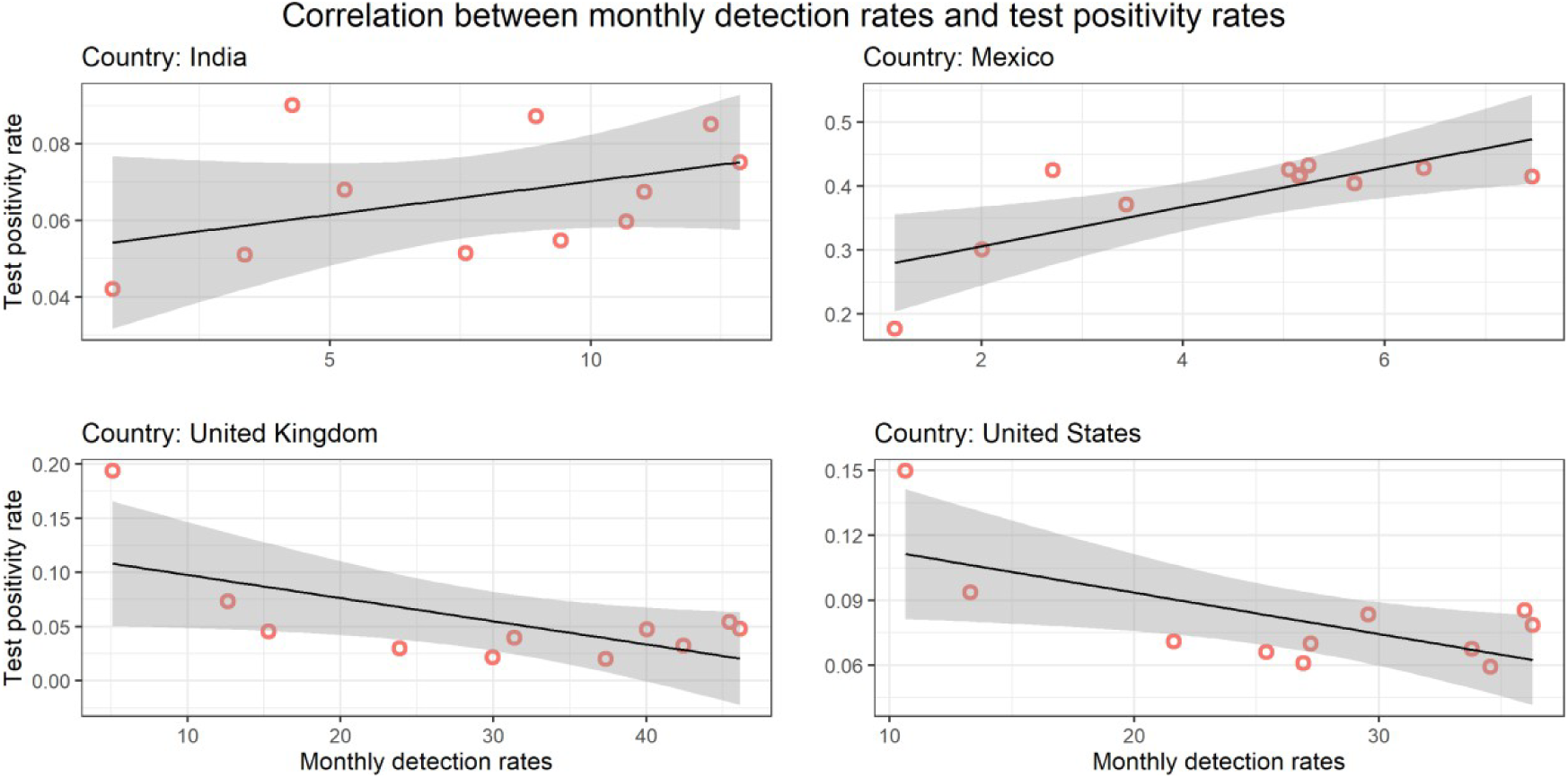
Correlation between detection rates and test positivity rates.

## DISCUSSION

This study provides an updated estimation of the true scale of the COVID-19 pandemic as of February 28, 2021, demonstrating that official case counts significantly underestimate the true number of infections. While testing infrastructure and knowledge of viral transmission improved over the first year of the pandemic, infections continued to rise. Our findings suggest that the cumulative number of infections in India, Mexico, the United Kingdom, and the United States may be more than six times the officially reported figures, highlighting the extent of underreporting.

A key strength of this study lies in its application of a country-specific probabilistic bias analysis model, rather than a broad global approach. By incorporating infection waves, viral mutations, and changing test positivity rates, this model adjusts for biases in reported data and provides a more realistic estimate of infection prevalence. The model also accounts for multiple testing and changes in detection rates over time, ensuring a dynamic assessment of pandemic progression in different settings. The ability to track infections over a one-year period allows for a comparative analysis of trends within and across countries, offering valuable insights into differences in surveillance effectiveness.

Our results indicate substantial disparities in case detection rates among the studied countries. The United Kingdom and the United States exhibit a relatively strong alignment between confirmed cases and estimated infections, reflecting more comprehensive testing and surveillance systems. However, India and Mexico show significantly weaker correlations, likely due to lower testing rates and more selective testing strategies. Mexico, in particular, had the lowest detection rate of true infections, consistent with its persistently high test positivity rate and limited testing infrastructure. This pattern underscores the importance of widespread testing in accurately tracking disease burden and highlights the challenges faced by countries with resource constraints.

A notable finding of this study is the relationship between testing rates, test positivity rates, and detection rates. Higher testing rates are associated with greater detection of infections, as evidenced by the United Kingdom’s and the United States’ relatively high detection rates. Conversely, limited testing capacity results in underreporting, as seen in Mexico, where the test positivity rate remained around 40% for an extended period. This aligns with previous research showing a strong inverse correlation between detection rates and test positivity, emphasizing the importance of scaling up testing efforts to improve surveillance accuracy.

The intuitive relationship between testing and detection is further supported by correlation analysis, which demonstrates statistically significant positive associations between testing rates and detection rates in all four countries. In the United Kingdom and the United States, a significant negative correlation is observed between test positivity and detection rates, confirming that expanded testing leads to a greater share of asymptomatic and mild cases being identified. However, Mexico presents an anomaly, where a weak positive correlation between test positivity and detection rate suggests ongoing data limitations or reporting inconsistencies. Further research is necessary to explore whether similar patterns exist in other low-testing settings.

Our findings are consistent with estimates from other modeling approaches and seroprevalence studies. Previous research using backcasting methods estimated that by September 2020, approximately 10% of the populations of Mexico, the United Kingdom, and the United States had been infected, with a lower prevalence in India (Noh and Daunser, 2021). Our model estimates similar trends, with slight variations likely attributable to differences in methodology. Other studies have estimated a detection rate of approximately 24% in the United States by late 2020, comparable to our model’s estimated detection rate of 27% (Pei et al, 2021; Phipps, Grafton, and Kompas, 2020). These similarities reinforce the robustness of our approach and suggest that the true burden of COVID-19 has been consistently underestimated across various methodologies.

Seroprevalence studies further validate our findings. Large-scale surveys conducted in India, Mexico, the United Kingdom, and the United States report seropositivity rates that align closely with our estimated infection rates. For instance, the Indian Council for Medical Research’s nationwide serosurvey estimated an infection rate of 6.6% by September 2020, closely matching our model’s estimate of 6.2% (Murhekar et al., 2021). Similar consistency is observed in Mexico, where our infection estimates align with seroprevalence data collected from urban populations (Rodríguez-Vidales et al., 2021). These comparisons suggest that our model provides a reasonable approximation of true infection rates, even in the absence of widespread serological data.

Despite its strengths, this study has several limitations. The model relies on national-level statistics, which may obscure regional variations in infection rates. This is particularly relevant for large, geographically diverse countries such as India, Mexico, and the United States, where testing infrastructure and healthcare access vary widely across regions. Subnational analyses would provide a more granular understanding of disparities in case detection and help identify localized hotspots of transmission.

Additionally, while the model indirectly accounts for factors such as mobility patterns and public health interventions by incorporating country- and time-specific data, it does not explicitly model these effects. The influence of lockdowns, mask mandates, and vaccination efforts on transmission dynamics remains an important area for future research. Estimating infections on a monthly basis also introduces some limitations, as intra-month variations in testing and case reporting may not be fully captured.

Extending this model beyond February 2021 would require adjustments to account for vaccine-induced immunity. While a straightforward approach would be to exclude vaccinated individuals from the susceptible population, this method poses challenges due to the lack of precise data on individuals who acquired immunity through prior infection. Many individuals recover from COVID-19 without a documented positive test result, making it difficult to separate those with natural immunity from those vaccinated. Future iterations of this model should incorporate improved surveillance data that track both prior infections and vaccination status.

Importantly, this study refrains from estimating the infection fatality rate (IFR) due to limitations in death reporting and excess mortality estimates. Many countries, including those studied here, have reported substantial excess deaths that exceed official COVID-19 mortality figures. Without reliable age-stratified mortality data, any IFR estimates would likely be biased and misleading. Further research is needed to reconcile reported deaths with estimated infection counts to provide a more accurate assessment of COVID-19’s true mortality burden.

The findings of this study underscore the urgent need for improved population health surveillance. Existing test-based case counts significantly underestimate true infections, particularly in settings with limited diagnostic capacity. To enhance future pandemic preparedness, we propose the integration of model-based infection estimates into public health reporting systems. Combining these estimates with seroprevalence data could provide a more comprehensive understanding of disease burden, guiding resource allocation and policy interventions.

Additionally, genomic surveillance must be expanded to track emerging SARS-CoV-2 variants effectively. While genomic sequencing has been widely implemented in high-income countries, many low-income settings lack the infrastructure for widespread variant tracking. Strengthening genomic surveillance in resource-limited regions is crucial to preventing the global spread of more transmissible or immune-evasive variants. A potential strategy is to prioritize sequencing in regions experiencing unexplained surges in infections, allowing for early detection and containment of new variants.

Herd immunity has been proposed as a long-term strategy for managing COVID-19, but our findings suggest that achieving national-level herd immunity may be insufficient. Viral transmission is often concentrated in densely populated urban areas, meaning that even if a country reaches an overall threshold for herd immunity, unprotected pockets of the population may still serve as reservoirs for transmission. This risk is especially pronounced in large, heterogeneous countries where regional vaccination rates vary widely.

A more nuanced approach involves the concept of an effective herd immunity threshold (eHIT), which accounts for ongoing public health measures and changes in viral transmissibility. For example, if 30% of a population has acquired immunity, maintaining an effective reproduction rate (Rt) below 1.43 would be necessary to prevent sustained transmission. Policymakers should consider such dynamic thresholds when designing mitigation strategies, rather than relying solely on static herd immunity estimates.

Ultimately, the findings of this study reinforce the importance of a sustained, coordinated global response to COVID-19. While vaccination efforts have made significant progress, the continued spread of new variants highlights the need for complementary measures such as widespread testing, genomic surveillance, and targeted public health interventions. The history of infectious disease eradication, such as smallpox, demonstrates that successful containment requires a multifaceted approach that combines vaccination with active case detection and community engagement.

To effectively manage COVID-19 and future pandemics, public health strategies must be adaptive, data-driven, and globally coordinated. A reliance on confirmed case counts alone is insufficient - integrating robust estimation models with seroprevalence studies and genomic tracking can provide a more comprehensive understanding of disease dynamics. Strengthening these surveillance mechanisms will be critical in ensuring an equitable and effective pandemic response worldwide.

## CONCLUSION

This study quantifies the true health burden of COVID-19 as of February 28, 2021, in four of the most affected countries by optimizing a probabilistic bias analysis model. The derivation of infection trends over time and across countries provides valuable insights into the first global pandemic of the 21st century. Our findings indicate that reported case numbers significantly underestimate the true scale of infections, with the pandemic spreading at an unprecedented rate. Within a year, cumulative infections in these four countries exceeded 200 million, with novel variants and mutations posing ongoing threats amid an imperfect vaccine rollout.

Although the most acute phase of the pandemic has now passed, understanding the true scale of infections during the pre-vaccination period remains vital. By reconstructing infection rates before widespread vaccine uptake, this study contributes to a more accurate historical record of the pandemic’s early trajectory, when surveillance systems were overwhelmed and case detection was highly inconsistent. These revised estimates offer an essential baseline for interpreting subsequent waves, assessing the impact of vaccination, and understanding the evolution of transmission dynamics.

Moreover, accurate retrospective infection burden estimates continue to have relevance today. They help quantify the long-term consequences of COVID-19, including post-acute sequelae, inform health-system planning, and strengthen preparedness for future epidemics by identifying weaknesses in early detection and reporting mechanisms. They also support ongoing epidemiological and economic analyses that depend on credible estimates of true infections rather than reported case counts.

## Data Availability

All data produced are available online at https://ourworldindata.org/coronavirus

