## Supplement for "Reassessing COVID-19’s First Year: Estimating Infections and Tracking Pandemic Trends with Probabilistic Bias Analysis"

Harsh Vivek Harkare

The aim of this research was to estimate the total number of cumulative SARS-CoV-2 infections in India, Mexico, the United Kingdom, and the United States using a probabilistic bias analysis method. The infections were estimated for a period from March 1, 2020 to February 28, 2021 with the intention of tracking the progression of the pandemic within one year. Patterns of movement in variables such as detection rates and population infection rate are observed to provide an insight into the infection trends in the selected group of countries. Correlations between the testing rate and detection rate of true infections are also observed.

### Reference study

My study builds upon the empirical approach implemented by Wu et al. estimating SARS- CoV-2 infections in the United States (Wu et al., 2020). The reference study procures data on US states and territories from the COVID Tracking Project from February 28, 2020 to April 18, 2020. The aim of the study was to estimate SARS-CoV-2 infections in the United States using probabilistic bias analysis to correct for biases due to incomplete testing and imperfect test accuracy during the aforementioned 51-day time period. Prior distributions of testing probabilities were defined for the entire period using available literature. Through Monte Carlo simulations, the uncertainty in testing probabilities was addressed to arrive at the estimated number of total infections. Prior distributions were defined for the 7 testing-related probabilities of P(*S*1|tested), P(*S*1|untested), P(*S*0|test +), *α*, *β*, test sensitivity and specificity. Finally, the reference model relates the number of untested individuals to the testing probabilities in order to estimate the true number of cumulative infections.

The reference study made 2 main assumptions related to the testing of individuals - (1) All test results in the obtained dataset were conducted using PCR-testing, and (2) a majority of the samples tested were collected with nasopharyngeal swabs. Additionally, the study assumes that the prior distributions are uncorrelated between states. The test positivity ratio is defined as a point estimate of the average probability of P(test + |tested) over the entire 51-day period. This was done to avoid unstable test positivity rates that would arise from very low testing rates in the beginning of the study period. In doing so, the estimation model essentially leaves out any potential fluctuations, however small, in infection trends during the study period. The study presents a national- and state-level count of the estimated total number of true SARS-CoV-2

infections in the United States along with the proportion of infections attributable to incomplete testing and imperfect test accuracy.

For the study, the general framework of the reference study’s estimation model has been borrowed, including the prior distribution definitions and some of the estimation formulae. Changes and additions to the model were made in the following areas:

- Enhanced modelling approach to consider following aspects:
  - Variation in monthly test positivity rates.
  - Infection waves in each country.
  - Effects of variants and mutations.
- Updated primary estimation formulae to account for the possibility of multiple testing of individuals to allow for a longer estimation period.
- Extended study period and range of countries.

The choice of countries included in this study puts a focus on the countries with some of the highest reported COVID-19 caseloads in the world. Estimating the true number of infections in these countries allows for the possibility of identifying and comparing infection trends over time. Additionally, the differences in their socio-economic characteristics such as income- levels and age-structures can potentially offer further insights into understanding the observed trends.

The empirical approach of the reference paper as well as my contributions to it have been described in further detail in the following sections.

#### Estimation of cumulative infections

For estimating the total number of cumulative SARS-CoV-2 infections in the countries of India, Mexico, the United Kingdom, and the United States, I employ the method of probabilistic bias analysis (Lash, Fox, and Fink, 2009).

Within the realm of quantitative bias analysis, a probabilistic bias analysis can be implemented to model greater uncertainty. Simple bias analysis suffers from the assumption that its bias parameters are fixed at values that confer no bias, which is difficult to confirm in a dynamic novel-pathogen setting such as the COVID-19 pandemic. On the other hand, the probabilistic bias analysis approach defines its bias parameters as random variables within defined probability distributions, and via repeated sampling from these distributions presents a bias- corrected frequency distribution of the desired estimates. Probabilistic bias analysis is a semi- Bayesian approach as it defines prior distributions for some of its bias parameters. Even when the probabilistic bias analysis approach does not use a formal likelihood function to model the relation between these prior distributions and the data, it has been shown to perform equally as well as Bayesian approaches in most instances (Lash, Fox, and Fink, 2009, p.179; MacLehose and Gustafson, 2012, p.156). A final argument in favour of the probabilistic bias analysis approach is that it is easier to use when compared to Bayesian modelling.

Given the uncertain nature of the COVID-19 pandemic, changing and constantly updating literature, and inherent uncertainty in methods of measurement, it is often the case that data to inform the bias parameters is insufficient. Even when there is information available to inform a certain prior, there is rarely enough evidence to completely disallow values above or below the mentioned range. The probabilistic bias analysis is employed to correct for this information bias. In this context, the choice of a beta distribution to model the prior distributions of bias parameters is appropriate. It provides a probability density function that is well-suited for assignment to proportions - such as sensitivity, specificity, or predictive or probabilistic values - since it is defined over the interval [0,1]. In choosing the beta distribution to model the bias parameters, the distribution was kept inclusive of a wide range of uncertainty. This is especially true towards the tails of the distribution, thus ensuring that most of the uncertainty regarding the testing probabilities is captured in the model. The parameters of a beta distribution are *α* and *β*.

$$E\left( X \right)= \frac{\alpha}{\alpha+ \beta}$$

$$Var\left( X \right)= \frac{\alpha\beta}{{(\alpha+ \beta)}^{2} (\alpha+ \beta-1)}$$

The reporting and documenting of the probabilistic bias analysis as an estimation approach is undertaken in accordance with the presentation guidelines reported in the book “Applying Quantitative Bias Analysis to Epidemiologic Data” (Lash, Fox, and Fink, 2009, p.175).

Prior distributions of testing probabilities were informed using a meta-analysis of relevant time- and country-specific literature on testing probabilities, clinical characteristics of SARS- CoV-2, and diagnostic accuracy of the testing equipment used, when available. These were defined for individuals based on the display of moderate-to-severe symptoms or mild-to-no symptoms. In accordance with CDC guidelines, moderate-to-severe symptoms are those requiring urgent medical attention or hospitalization. Individuals with dyspnea (shortness of breath), chest pain, confusion, inability to stay awake, and hypoxemia are directed to seek medical care immediately (CDC, 2021c). Additionally, severe forms of any other symptoms such as high fever or persistent dry cough are also treated as severe symptoms of COVID-19. *S*1 is defined to act as an indicator of moderate-to-severe symptoms of COVID-19, while *S*0 indicates mild-to-asymptomatic infections.

Utmost effort has been made to inform the priors for each of the individual countries with time- and country-specific literature but given the dynamic nature of the pandemic as well as the limited number of data and studies available on SARS-CoV-2 testing probabilities, some priors for later time periods have been informed based on studies conducted within earlier time periods as well. All parameters of the prior distributions including the mean and variance have accordingly been informed by the available literature.

The discovery of viral mutations of SARS-CoV-2 and their effect on the epidemiological characteristics of COVID-19 have also been incorporated in my model. In the absence of country-specific literature on the impact of different SARS-CoV-2 variants on each of the testing probabilities, the priors have been updated based on the documented literature on the increased transmissibility and infectivity of the SARS-CoV-2 variants (Davies et al., 2021a, p.270; Gordon et al., 2020, p.13; Thorne et al., 2021, p.15; Zhang et al., 2020, p.2). Detailed descriptions of each prior distribution definition can be found in section 4.7 - “Defining infection waves and prior distributions.”

My estimation model quantifies the uncertainty in testing probabilities by randomly sampling from each prior distribution 10^4^ times. The final number of estimated true infections of SARS- CoV-2 is given by a formula relating the number of susceptible individuals to the testing probabilities. I report the median of the Monte Carlo distribution of infection estimates in each country along with the 2.5^th^ and 97.5^th^ simulation interval. The reported estimated number of infections are inclusive of both confirmed COVID-19 cases and undiagnosed infections.

Since the efficacy and accuracy of the results of a probabilistic bias analysis method are heavily dependent on the chosen priors, documentation on informing the priors becomes especially important. In consequence, the approach towards defining and informing the prior distributions is evidence-based and has been purposefully kept transparent, clear, and well-documented. Given that the SARS-CoV-2 virus is a novel coronavirus strain that is constantly mutating in countries around the world, the prior distributions have been defined with a greater uncertainty margin to account for this dynamic nature.

To effectively estimate the true number of SARS-CoV-2 infections over time in the chosen sample countries, I make the following assumptions in my model:

1) Since a majority of the tests conducted in the sample countries were tested via the RT- PCR method, it has been assumed that all tests have been conducted via RT-PCR testing using nasopharyngeal swabs to generalize the prior definitions for test sensitivity and specificity.
2) In addition, testing is assumed to be conducted within the first two weeks of infection.
3) Test specificity of RT-PCR testing is assumed to be unaffected by SARS-CoV-2 variants.
4) Past infection from SARS-CoV-2 has been assumed to provide lasting immunity.

#### General definitions of the prior distributions

**P(*S*1|tested)** : The distribution of this prior is defined as a truncated beta distribution and is indicative of the share of moderate-to-severe symptomatic cases among those being tested. During the peak of a pandemic, this prior would be higher as with a surge in infections, testing capacity is stretched to a limit, and only the more severe cases are tested and diagnosed. This is also true for the early months of the pandemic, when testing capacities were still being developed and the limited available testing equipment were prioritised for cases with relatively severe, more visible symptoms (CDC, 2021b).

**P(*S*1|untested)** : Indicative of the suspected spread of moderate-to-severe symptomatic cases of COVID-19 in the general population, the P(*S*1|untested) prior is defined as a truncated beta distribution. As with P(*S*1|tested), this prior distribution would also be expected to be higher during the peak of a pandemic as the spread of the virus within a population grows and undetected infections rise.

***α*** : Defined as P(test + |*S*1, untested) divided by the empirical test positivity rate, *α* relates the probability of an undiagnosed individual exhibiting moderate-to-severe symptoms to the empirical estimate P(test + |tested). The prior *α* is modelled to vary every month starting May 1, 2020 and is sensitive to the dynamic nature of a pandemic by capturing the surge and fall of infections. The prior distribution is defined as a truncated beta distribution. Most studies assessed to inform this prior are, in fact, used to inform P(test + |*S*1, untested), which is the probability of an untested individual exhibiting moderate-to-severe symptoms testing positive for the novel coronavirus infection.

$$\alpha=\frac{P(test +| S_{1}, untested)}{P(test+| tested)}$$

***β*** : Similar to *α*, *β* is defined as P(test + |*S*0, untested) divided by the empirical test positivity rate and relates the probability of an undiagnosed individual exhibiting mild-to-no symptoms to the empirical estimate P(test + |tested). *β* is modelled to vary each month starting May 1, 2020 and is sensitive to the peaks and troughs in infection rates. It is defined as a truncated beta distribution. Instead of directly informing the prior *β*, the current available literature was used to inform P(test + |*S*0, untested), which is the probability of an untested individual exhibiting mild-to-no symptoms testing positive for a SARS-CoV-2 infection.

$$\beta=\frac{P(test +| S_{0}, untested)}{P(test+| tested)}$$

**P(*S*0|test +)** : The distribution of P(*S*0|test +) is defined as a truncated beta distribution. It is indicative of the proportion of asymptomatic to mild-symptomatic cases of SARS-CoV-2 infections being detected.

**Test sensitivity (*S*e)** : Test sensitivity is defined as the ability of a particular test to correctly identify those with a disease or an infection. The distribution of SARS-CoV-2 test sensitivity is defined as a truncated beta distribution. Since testing equipment varies from country-to-

country, this prior is informed by literature from individual countries based on the most common testing methods employed there.

**Test specificity (*S*p)** : Defined as the ability of a test to correctly detect the absence of a disease, the prior distribution of the test specificity for SARS-CoV-2 is defined as a truncated beta distribution. Similar to test sensitivity, this prior is also informed by literature from different countries based on the testing methods in use in those particular countries.

Correctly setting the value of sensitivity and specificity is important as it has many implications on the overall case numbers. It also influences the isolatory tendencies of individuals, as individuals with negative test results are less likely to self-isolate and practice social distancing. It should be noted that the timing of the test administration can impact the effectiveness of its results. Test sensitivity has been found to be highest soon after symptom onset after which it declines and is close to zero more than 35 days after symptom onset (Xiao et al., 2020, p.2). Serological test sensitivity, in contrast, increases with duration from the date of symptom onset (Miller et al., 2020, p.13881). In accordance with the reference paper, the prior for test sensitivity builds on the assumption that testing was conducted within the first 2 weeks of being infected with SARS-CoV-2 when test sensitivity is maximized.

Additionally, SARS-CoV-2 variants and mutations have also been documented to limit testing efficacy. A study looking at the genomic sequencing of different SARS-CoV-2 mutations established that most diagnostic assays for detecting a SARS-CoV-2 infection are not designed to test a plurality of gene primers and hence are negatively affected by the variants in terms of test sensitivity (Wang et al., 2020, p.5207). Another genomic study discovered that any changes to the genomic structure of the novel coronavirus can negatively affect its detection by RT- PCR tests, since not all diagnostic assays include multiple genetic targets for detecting the virus (Tahan et al., 2021, p.4). Accordingly, prior distributions defining the test sensitivity for the third wave of all countries have been corrected downwards. Test specificity is assumed to not be affected.

#### Sampling the priors

For estimating the cumulative number of SARS-CoV-2 infections and tracking the progression of the COVID-19 pandemic, yearlong data from a period of March 1, 2020 to February 28, 2021 was used. Contrary to the reference paper (Wu et al., 2020), I chose to allow the test positivity rate to vary over time to capture the changes in infection trends over time. For each country, the test positivity rate is calculated as the cumulative number of new confirmed cases detected in a particular month divided by the cumulative number of new tests conducted in that same month, beginning on the 1st until the end of the month. This empirical estimate P(test +

| tested) is therefore a series of point estimates of the average test probabilities over each month. Since at the beginning of the pandemic testing capacities were very low in the studied countries and because testing data was not easily available, P(test + | tested) was calculated as a point average over two months starting from March 01, 2020 until April 30, 2020 in a similar manner as described above.

To calculate monthly infection estimates, I run a probabilistic bias analysis for the two months of March and April 2020 starting March 01, 2020, and then for each month until February 28, 2021. For each monthly estimate, only the cases and tests reported in that particular month are taken into consideration. An advantage of this method over the reference model is that the monthly test positivity rates are better aligned with the fluctuations in infection trends during pandemic waves, and are more representative of the size and scale of viral outbreaks in a particular month. This allows for a more time- and situation-sensitive modelling of the estimated infections.

For the two months in the beginning of the pandemic (March 01, 2020 to April 30, 2020) and for each month thereafter, 10^4^ values were sampled from the distributions of P(*S*1 | tested), P(*S*1 | untested), *α*, *β*, test sensitivity, and test specificity for each country. The probabilities of an untested individual exhibiting moderate-to-severe or mild-to-no symptoms testing positive on their next test, P(test + | *S*1, untested) and P(test + | *S*0, untested) respectively, are sampled from the empirical estimate of test positivity P(test + | tested) and *α*, *β*.

#### Constraining of prior distributions

The prior distribution of P(*S*0 |test +) has been informed by applying the method of Bayesian melding to incorporate information on the prior from two different sources: (1) Scientific evidence from population-representative studies, and (2) via a function of the parameters P(*S*1 | untested), *α*, and *β*. In doing so, P(*S*0 |test +) becomes a joint melded distribution constrained by its defining parameters. This makes the prior adhere closely to the observed data, making the probabilistic bias analysis equivalent to a Bayesian approach (MacLehose and Gustafson, 2012, p.156).

The function *φ* = P(S0 |test +) informs the prior distribution from available scientific evidence (defined below), while *θ* = {P(*S*1 | untested), *α*, *β*} is the function relating P(S0 |test +) with the other three prior distributions. The relationship between *φ* and *θ* is given by the function (*M*: *θ* → *φ*):

$$P(S_{0}|test +)= \frac{\beta(1-P(S_{1}|untested)}{\beta(1-P(S_{1}|untested))+ \alpha P(S_{1}| untested)}$$

Via Bayesian melding, information from the two functions has been combined to define a third function M which is a joint melded prior distribution on {*θ*, *φ*} sampled 10^5^ times (Poole and Raftery, 2000, p.1246). The function M is used to inform the final prior distribution of P(*S*0|test +) via the Sampling-Importance-Resampling algorithm (Poole and Raftery, 2000, p.1249).

#### Bias corrections

Sampled variates of the defined prior distributions were used as inputs to the estimation formulas (2) - (4). Formulas (2) and (3) are used to correct for incomplete testing and to quantify the number of undiagnosed SARS-CoV-2 infections in each country. Estimation formula (4) corrects for imperfect test accuracy in terms of imprecise test sensitivity and specificity, and returns *N*^*^, the estimated number of SARS-CoV-2 infections.

$$N_{untested, S_{1}}^{+}=P\left( S_{1}|untested \right)\times P\left( {test + |S}_{1}, untested \right)\times N_{susceptible}$$

$$N_{untested, S_{0}}^{+}=(1- P\left( S_{1}|untested \right)) \times P\left( {test + |S}_{0}, untested \right)\times N_{susceptible}$$

*N^+^*	is the estimated number of individuals with moderate or severe symptoms with undiagnosed infections who would have tested positive if complete and perfect periodical testing were made. *N^+^*	is the analogous value for undiagnosed infections with mild-to- no symptoms.

untested, *S*1

untested, *S*0

The resultant values from formulas (2) and (3) are added to the confirmed reported infections to get the total number of estimated infections uncorrected for imperfect testing biases (*N^+^* = *N^+^*	+ *N^+^*	+ *N^+^*). This value is then used in formula (4) correcting for the bias of imperfect testing (Diggle, 2011, p.1; Rothman, Greenland, and Lash, 2008; Wu et al., 2020, p.7).

untested, *S*1	untested, *S*0	confirmed

$$N^{*}=\frac{(N^{+}-\left( \left( 1- S_{p} \right)\times N \right))}{S_{e}+S_{p}-1}$$

Instead of using the number of individuals susceptible to infection (*N*susceptible) to quantify the number of missed infections, the reference paper (Wu et al., 2020) uses the number of untested individuals (*N*untested) in the estimation formulas (2) and (3). Consequently, the study makes an inherent assumption that all individuals who have been tested negative at any point over the 51-day study period will not be infected after testing and are thus left out of the estimation process. The validity of this assumption can be called into question even when estimating infections for a short period early in the pandemic timeline when community rates of transmission were low or moderate in the United States (Ritchie et al., 2020). When estimating infections for a longer time period, however, this assumption becomes ineffective as there is a higher chance of infection for every individual with passing time and rising infections, as was the case with the countries included in this study. As the number of people getting tested for COVID-19 - irrespective of the result - grows over the course of the pandemic, the number of untested individuals (*N*untested) would keep falling which would lead to an artificial suppression of the estimated number of infections. Additionally, all of the countries included in this study measure testing data as the number of tests administered instead of the number of individuals tested. A single individual getting tested multiple times would also falsely reduce the number of untested individuals (*N*untested) and consequently, the estimated number of true infections. Thus, a blind application of the reference model’s estimation approach would result in an artificially low number of estimated infections.

To overcome this limitation, I substitute the number of untested individuals (*N*untested) with the number of individuals still susceptible to an infection (*N*susceptible). These are defined as individuals with neither a confirmed infection test result nor estimated to be infected as per my model. For the period of March 1, 2020 to April 30, 2020, the number of susceptible individuals is defined by subtracting the number of reported COVID-19 cases from each country’s population (*N*susceptible = *N* - *N*^+^	). For each month thereafter, *N*susceptible is defined as a monthly value subtracting the number of infections; estimated and confirmed, until the end of the previous month from the country’s population (*N*susceptible, t = *N* - *N*^*^ ). In using *N*susceptible in contrast to *N*untested for estimating the total number of infections, my model allows for the possibility of individuals getting tested multiple times with negative test results and still being susceptible to a viral infection, which is a reality of the pandemic in many countries.

t - 1

confirmed

To effectively model for the estimated number of true infections using *N*susceptible, I assume zero reinfections. In other words, individuals who have already been infected with SARS-CoV-2 develop a lasting immunity and cannot get reinfected. Only these individuals are then left out of the correction formulae (2) and (3) when estimating the total number of infections. My assumption is based on scientific evidence that reinfection rates have been documented to be below 1%, and that immunity from a SARS-CoV-2 infection lasts for as long as 8 months after infection (Dan et al., 2021, p.594; Hansen et al., 2021, p.1207).

In accordance with the guidelines for conducting a probabilistic bias analysis, I repeated the entire above-described process 10^4^ times to fully account for the uncertainty in modelling the total number of estimated infections *N*^*^. Of the distribution of estimated infections calculated for each country, the median value along with the 2.5^th^ and 97.5^th^ quantile is presented.

I chose not to calculate the share of missed infections attributable to incomplete testing and imperfect test accuracy as with an increase in the true prevalence level, the relative impact of an imperfect test accuracy decreases, thereby attributing most missed infections to insufficient testing (Diggle, 2011, p.2). This is especially true for my studied period of one year with constantly rising SARS-CoV-2 infections.

#### Correlation analysis

From the estimated number of true SARS-CoV-2 infections, I derive cumulative monthly estimates of the implied detection rate of true infections for each country. To observe the intuitively-linear relationship between detection rates and testing rates, I conduct a correlation analysis using the Pearson’s coefficient correlation method. Since the observations are not identically distributed across countries, I conduct the correlation analysis for each individual country. The correlations are conducted between the monthly detection rates of true infections and testing rates such as average daily tests conducted per 1,000 and the test positivity rate.

#### Defining infection waves and prior distributions

Literature to inform priors has been taken from a variety of sources from a range of countries. Resources such as the Coronavirus Disease 2019 Associated Hospitalization Surveillance Network (COVID-NET) have also been utilized. COVID-NET is a population-based surveillance system that collects data on laboratory-confirmed COVID-19 associated hospitalizations in a network of over 250 acute-care hospitals in 14 states in the United States. Roughly 10% of the country’s population is covered (CDC, 2020c).

For each of the four countries included in this study, the priors have been defined for three sets of time periods of 4 months each and will hereafter be referred to as ‘waves’. The timeframe of the first, second, and third wave for each country is defined from March 1, 2020 to June 30, 2020, July 1, 2020 to October 31, 2020, and November 1, 2020 to February 28, 2021 respectively. The timeframes have been generalized and chosen to best fit the observed trends of reported infections in each country. To account for extremities in infection trends within a single wave, the variance of the defined prior distributions has been fixed at a higher level for waves with drastically fluctuating confirmed infection rates. Within the model, the first wave is defined to mirror the beginning of the pandemic, the second the middle, and the third wave is representative of when most countries were affected by the mutations and variants of concern of the coronavirus. To run the probabilistic bias analysis for monthly estimates of true infections, prior distributions defined for one of the three infection waves are selected. Overall, 12 sets of priors have been created for four countries to model the estimated infections based on the peaks and troughs of the COVID-19 pandemic.

Below, I tabulate the country-wise definitions of prior distributions and present the literature used to inform them.

##### India

| Prior | Wave 1 | Wave 2 | Wave 3 |
| --- | --- | --- | --- |
| P(*S*1\|tested) | 0.75 - 1 (mean: 0.9) | 0.7 - 1 (0.9) | 0.5 - 1 (0.8) |
| P(*S*1\|untested) | 0.001 - 0.1 (0.02) | 0.001 - 0.13 (0.05) | 0.001 - 0.2 (0.1) |
| *α* | 0.2 - 0.6 (0.25) | 0.4 - 0.7 (0.475) | 0.4 - 0.85 (0.65) |
| *β* | 0.01 - 0.3 (0.065) | 0.025 - 0.4 (0.125) | 0.02 - 0.35 (0.15) |
| P(*S*0\|test +) | 0.45 - 0.95 (0.8) | 0.15 - 0.85 (0.5) | 0.2 - 0.6 (0.4) |
| Test sensitivity (*S*e) | 0.82 - 1 (0.975) | 0.82 - 1 (0.975) | 0.8 - 1 (0.95) |
| Test specificity (*S*p) | 0.9998 - 1 (0.99995) | 0.9998 - 1 (0.99995) | 0.9999 - 1 (0.99995) |

**Table S1:** Definitions of prior distributions for India

###### P(S1|tested)

An Indian study testing 18,600 individuals with COVID-19 symptoms, travel history to an affected region, or healthcare workers (HCWs) based on the recommendations issued by the Department of Health Research found that 64.5% of the participants exhibited influenza-like illness symptoms (Yadav et al., 2021, p.3). A study documenting the clinical profiles of 114 patients admitted to a tertiary-care hospital in Chandigarh, India during April-May 2020 found 77% of the participants to have a fever, 21% had symptoms of cough, and 17% had hypoxia. Overall, 21% of all participants required intensive care or ventilation, or died (Soni et al., 2021, p.120). Between May 11 and June 28, 2020, the All India Institute of Medical Sciences (AIIMS) hospital in New Delhi screened and collected data on 235 patients confirmed to have COVID-19. Of these, 81.7% had moderate, severe, or critical illnesses (Kayina et al., 2020, p.101). A study testing 3745 individuals at a designated COVID-19 hospital in New Delhi from April through September 2020 found 68% to have the symptoms of dry cough, dyspnea, or chest pain (Saxena et al., 2021, p.2489).

###### P(S1|untested)

A study testing HCWs at a hospital in Mumbai, India for a period of 4 months from April to August 2020 found 9.5% of the participants to be symptomatic. Less than 1% had to be hospitalized for critical care (*N* = 3711) (Mahajan et al., 2020, p.19). Another study analysing surveillance data from the Indian Council of Medical Research (ICMR) from January to April 2020 found only 1% of all participants to have severe symptoms (*N* = 1,021,518) (Abraham et al., 2020, p.432). A seroprevalence study conducted in both slum and non-slum communities of Mumbai, India discovered the seroprevalence positivity to be 54% and 16% respectively, ahigh percentage of which are deemed to be asymptomatic infections (Malani et al., 2020, p.6). It should be noted that Mumbai is a major financial hub for the country and is very densely- packed. Results from the city therefore cannot be considered directly representative for the whole country. Although not representative of the general population, evidence from a maternity hospital testing all pregnant women admitted for delivery (*N* = 3,165) in September- November 2020 shows that 9.4% of the participants exhibited symptoms (Gupta, Kumar, and Sharma, 2021, p.3).

##### α

A retrospective case-series study of 82 hospitalized patients with severe respiratory illnesses in New Delhi in April 2020 found 39% of the patients to be SARS-Cov-2 positive (A. Aggarwal et al., 2020, p.22). Another study based in Jaipur, India, tested 501 individuals suspected of having COVID-19 in April 2020. Of all participants exhibiting severe symptoms, 93% were confirmed positive (Sharma et al., 2020, p.5). A study conducted in New Delhi in April 2020 first collected information on symptoms from HCWs working in hospitals around Delhi. Test positivity among those exhibiting symptoms was documented at 5.2% (Jha et al., 2020, p.119). An Indian study based in Mumbai screened 18,600 people for COVID-19 during the country’s first outbreak from March to June 2020. Of the 3,529 individuals displaying severe flu-like symptoms, 65.9% tested positive for the virus (Yadav et al., 2021, p.3). A study collected data on 3165 pregnant women from a maternity hospital in North India between September and November 2020 to find test positivity rates among symptomatic women to be 5.1% (Gupta, Kumar, and Sharma, 2021, p.4).

##### β

A Jaipur-based study testing 501 individuals suspected of having COVID-19 found 49.5% of participants exhibiting mild or no symptoms to be positive (Sharma et al., 2020, p.5). Test positivity among those exhibiting no symptoms was documented at 1.1% in a study conducted in New Delhi testing HCWs in April 2020 (Jha et al., 2020, p.119). A COVID-19 screening study conducted in Mumbai between March-June 2020 found that of 8,703 participants exhibiting mild or no symptoms, 25.9% tested positive. When including participants exhibiting ‘other’ symptoms, this number goes up to 34.5% (Yadav et al., 2021, p.3). A COVID-19 screening at a maternity hospital found test positivity rates among asymptomatic participants to be 3.2% (Gupta, Kumar, and Sharma, 2021, p.3).

###### P(S0|test +)

A data-surveillance study covering pan-India testing between January to April 2020 documented 16% of all positive SARS-CoV-2 infections to be mild-symptomatic. A further 19.6% were asymptomatic (Abraham et al., 2020, p.432). An Indian study looking at the clinical outcomes of individuals with asymptomatic or mild SARS-CoV-2 infections tested 1,263 patients to find 89.4% with non-severe symptoms or outcomes in May 2020 (Krishnasamy et al., 2020, p.87). Of 413 HCWs confirmed positive at a hospital in Mumbai, 15% were asymptomatic (Mahajan et al., 2020, p.19). Another Mumbai-based COVID-19 screening study found 24% of all participants with a confirmed SARS-CoV-2 infection to have asymptomatic infections. A further 37% had mild symptoms (Yadav et al., 2021, p.3). Of 108 pregnant women confirmed positive for COVID-19, 86% were found to be asymptomatic between September-November 2020 by a study based in Jammu and Kashmir, India (Gupta, Kumar, and Sharma, 2021, p.4).

###### Test sensitivity (Se)

An evaluation of the diagnostic accuracy of RT-PCR assays at the AIIMS in New Delhi collected a total of 990 nasal and throat swabs to ascertain the test sensitivity at 81.8%. In patients who are tested within 5 days of symptom onset, the sensitivity is higher at 85.9% (Gupta et al., 2021, p.129). Another study documented the use of RT-PCR test kits by Meril Diagnostics which have an established sensitivity of 100% (95% CI 83-100%) (Gupta, Kumar, and Sharma, 2021, p.3; Meril Diagnostics, 2020).

###### Test specificity (Sp)

A study evaluating the diagnostic accuracy of rRT-PCR assays in New Delhi, India established the test specificity to be 99.6% after collecting both throat and nasal swabs from 330 individuals (Gupta et al., 2021, p.129). Test specificity of RT-PCR assays was established at 100% by a study in North India (Gupta, Kumar, and Sharma, 2021, p.3; Meril Diagnostics, 2020).

##### Mexico

| Prior | Wave 1 | Wave 2 | Wave 3 |
| --- | --- | --- | --- |
| P(*S*1\|tested) | 0.8 - 1 (0.9) | 0.85 - 1 (0.925) | 0.75 - 1 (0.875) |
| P(*S*1\|untested) | 0 - 0.1 (0.06) | 0 - 0.15 (0.06) | 0 - 0.21 (0.8) |
| *α* | 0.3 - 0.6 (0.35) | 0.25 - 0.5 (0.3) | 0.4 - 0.8 (0.5) |
| *β* | 0 - 0.2 (0.06) | 0 - 0.2 (0.05) | 0.05 - 0.3 (0.15) |
| P(*S*0\|test+) | 0.4 - 0.8 (0.6) | 0.4 - 0.9 (0.65) | 0.4 - 0.8 (0.5) |
| Test sensitivity (*S*e) | 0.85 - 1 (0.9) | 0.85 - 1 (0.9) | 0.8 - 1 (0.9) |
| Test specificity (*S*p) | 0.9998 - 1 (0.99995) | 0.9998 - 1 (0.99995) | 0.9998 - 1 (0.99995) |

**Table S2:** Definitions of prior distributions for Mexico

###### P(S1|tested)

Of the 309 patients tested at a tertiary-care center in Mexico City between February-April 2020, 80% had a fever, 41% dyspnea, and 38% chest pain (Ortiz-Brizuela et al., 2020, p.168). A cross-sectional study conducted in Mexico between June-July 2020 tested 100 individuals to find 47% having severe symptoms (Martinez-Fierro et al., 2021a, p.18). Testing 35,059 HCWs in Mexico City until July 2020 led to the discovery of 11,226 SARS-CoV-2 infections. 14% of all tested participants had one of the severe outcomes of hospitalization, pneumonia, or mechanical ventilation. A further 1% died (Antonio-Villa et al., 2020, p.18).

###### P(S1|untested)

A population-based screening study conducted from April to August 2020 found 5.8% of all participants to have moderate to severe symptoms (*N* = 1279) (Martinez-Fierro et al., 2021b, p.5). From a study conducted in Zacatecas, Mexico, in June-July 2020, around 8-9% of the participants with a documented contact with a SARS-CoV-2 positive individual exhibited symptoms of headache or fever (Martinez-Fierro et al., 2021a, p.18). COVID-19 testing data collected from June through September 2020 across 26 Mexican states on 482,413 individuals showed the prevalence of symptoms among the general population. 8.1% had abdominal pain, 14.4% suffered from dyspnea, and 22% ran a fever (Fernández-Rojas et al., 2021, p.576). A cross-sectional study conducting universal screening of pregnant women admitted to a tertiary- care facility in Mexico City found 2.7% of all patients to have symptomatic SARS-CoV-2 infections (*N* = 1880) (Hernández-Cruz et al., 2021).

##### α

A study analyzed data from the National Epidemiological Surveillance System in the greater Mexico City area. With data collected on 296,157 patients until 1 August 2020, it was found that 36% of the participants with respiratory symptoms of dyspnea had tested positive for COVID-19 (Bello-Chavolla et al., 2021, p.656).

##### β

An observational study conducted in Mexico City in June 2020 tested 139 patients, all of whom were exhibiting mild-to-moderate symptoms of COVID-19. Test positivity among this sample was established at 51.7% (Romero-Gameros et al., 2020, p.987). Another study looking at data from the National Epidemiological Surveillance System in Mexico with data until September 2020 discovered the test positivity rate among patients with non-severe non-respiratory symptoms to be 22.9% (Bello-Chavolla et al., 2021, p.656).

###### P(S0|test +)

Universal screening of pregnant mothers at a tertiary care center in Mexico City in May 2020; at a time when community transmission was high, found 86% of the 70 SARS-CoV-2 positive participants to have asymptomatic infections (Cardona-Pérez et al., 2021, p.5). A Mexican study evaluating the role of close contacts in COVID-19 transmission conducted between June- July 2020 found 73.5% of all positive close-contact participants to have only non-severe symptoms for the duration of their illness (Martinez-Fierro et al., 2021a, p.16). A population- based screening of the Mexican population in Zacatecas from April-August 2020 found that 36% of the participants with a positive test result were asymptomatic. A further 6% showed only 1 symptom throughout their illness (Martinez-Fierro et al., 2021b, p.4). Another mass- testing study collected data from June-September 2020 and found that 14-50% of participants with a positive SARS-CoV-2 test exhibited various mild symptoms (Fernández-Rojas et al., 2021, p.576).

###### Test sensitivity (Se)

Testing in Mexico was carried out using RT-PCR assays developed by the CDC (Martinez- Fierro et al., 2021b, p.4). The CDC developed RT-PCR test has analytical sensitivity ≥95% for RNA concentrations ≥1 copy μL^−1^ (CDC, 2020b). Another study conducted in Mexico stated that RT-PCR test sensitivity is demonstrated at 95% (Lewis et al, 2020, p.834).

###### Test specificity (Sp)

The SARS-CoV-2 RT-PCR diagnostic panel developed by the CDC is designed to minimize the chance of a false positive, so test specificity can be assumed to be very high in laboratories that comply with standard best practices, such as the use of negative controls (CDC, 2020b).

##### United Kingdom

| Prior | Wave 1 | Wave 2 | Wave 3 |
| --- | --- | --- | --- |
| P(*S*1\|tested) | 0.7 - 1 (0.85) | 0.8 - 1 (0.9) | 0.6 - 1 (0.8) |
| P(*S*1\|untested) | 0.01 - 0.4 (0.07) | 0.01 - 0.4 (0.05) | 0.05 - 0.5 (0.1) |
| *α* | 0.4 - 0.65 (0.5) | 0.35 - 0.55 (0.425) | 0.5 - 0.75 (0.6) |
| *β* | 0.05 - 0.35 (0.15) | 0.04 - 0.3 (0.15) | 0.07 - 0.5 (0.2) |
| P(*S*0\|test+) | 0.3 - 0.7 (0.6) | 0.45 - 0.7 (0.65) | 0.25 - 0.5 (0.4) |
| Test sensitivity (*S*e) | 0.87 - 1 (0.94) | 0.87 - 1 (0.94) | 0.85 - 1 (0.93) |
| Test specificity (*S*p) | 0.9998 - 1 (0.99995) | 0.9998 - 1 (0.99995) | 0.9998 - 1 (0.99995) |

**Table S3:** Definitions of prior distributions for the United Kingdom

###### P(S1|tested)

A prospective-cohort study in 208 hospitals across the United Kingdom enrolled 20,133 patients between February and April 2020. 70% displayed symptoms of cough, fever, or dyspnea. 55% required assisted breathing, 26% required ventilation, and 17% were admitted to intensive care units. Overall, 26% of the participants died (Docherty et al., 2020a, p.3-6). Another study ascertaining clinical outcomes for patients admitted to 166 hospitals in the UK between February and April 2020 documented 69% of participants to exhibit symptoms of fever, cough, and shortness of breath (*N* = 16,749). Overall, 17% of the participants required long-term medical attention, and 33% died (Docherty et al., 2020b, p.8). The Office for National Statistics tested 9,018 individuals between October 2020 and January 2021 to discover 14.4% exhibiting severe flu-like symptoms (Office for National Statistics, 2021a).

###### P(S1|untested)

A COVID-19 screening program of HCWs at a maternity hospital in London found 29% of the participants to be symptomatic (Khalil et al., 2021, p.24). A community-prevalence survey study conducted in the United Kingdom in May 2020 discovered that 0.13% of the participants (*N* = 120,610) tested positive for SARS-CoV-2 when using a RT-PCR test. Of these, only 39 participants (0.03% of the base sample) reported having had symptoms (Riley et al., 2020, p.7).

Another antibody test conducted in the UK during June-July 2020 found the adjusted test- prevalence rate to be 6% (*N* = 109,076), and overall 3.1% of the sample population reported one or more symptoms of fever, persistent cough, and a loss of taste or smell (Ward et al., 2020, p.7). A meta-analysis of 37 nationally-representative surveys conducted between March 2, 2020 and January 27, 2021 collected information on 74,697 responses from 53,880 people living in the United Kingdom. Between May 26, 2020 and January 27, 2021, 51% of the respondents reported symptoms of fever, cough, and a loss of taste and smell. For the first week of April 2020, the number was reported at close to 70%. Similarly for the first week of May, 65% of respondents exhibited severe symptoms of COVID-19. In the last week of January 2021, the same number was 50.8%. It must be noted that these numbers are prone to a self- selection bias (Smith et al,, 2021, p.4).

##### α

Between April to August 2020, the United Kingdom Office for Statistics tested 262 individuals exhibiting symptoms of cough, fever, or a loss of taste or smell, and found 6.9% to be positive for COVID-19 (Office for National Statistics, 2021a). An antibody surveillance study conducted in the United Kingdom in the months prior to May 2020 found that 17% of the population in London and 5% elsewhere in the United Kingdom tested positive for SARS- CoV-2 antibodies (Burki, 2020, p.e63). In May-June 2020, the Office for National Statistics in the United Kingdom found 26.3% of individuals exhibiting the symptoms of cough, fever, or loss of taste to be confirmed positive for a SARS-CoV-2 infection. The share of the same had risen to 37% in September 2020 (Office for National Statistics, 2021a).

##### β

P(test + |*S*0, untested) is estimated to be 65.03% based on a study in the United Kingdom wherein 2,618,862 individuals self-reported their symptoms on a smartphone-app. The study compared individuals with a symptom of loss of smell or taste and found that 65% of positive cases had this symptom (Menni et al., 2020, p.1037). Mass-testing of HCWs in Sheffield, United Kingdom, in March 2020 found the test positivity rate to be 18% among mildly- symptomatic cases (*N* = 1533) (Keeley et al., 2020, p.2). Data from the Office for National Statistics in the UK shows that 42.1% of individuals exhibiting mild symptoms between May- June 2020 tested positive for the novel coronavirus. In August, the same number had risen to 57.7%. Between October 2020 and January 2021, 52.5% of participants with no symptoms tested positive. A further 35% exhibited only non-severe symptoms (Office for National Statistics, 2021a).

###### P(S0|test +)

Regular testing of HCWs at a London hospital over multiple weeks starting March 2020 found that the share of asymptomatic cases among those confirmed positive varied between 1.1% - 7.1% over a period of 5 weeks (Treibel et al., 2020, p.1609). Another population-representative study carried out in the UK between April 26, to November 1, 2020 enrolling 280,327 individuals found that 45-68% of positive infections reported no symptoms and remained that way (Pouwels et al., 2021, p.30).

###### Test sensitivity (Se)

As per the methodology of the COVID-19 infection survey conducted by The Office for National Statistics in the United Kingdom, COVID-19 test specificity is established to be between 85-98% (Office for National Statistics, 2021c, p.8).

###### Test specificity (Sp)

Test sensitivity of COVID-19 tests administered in the United Kingdom is established to be higher than 99.92% (Office for National Statistics, 2021c, p.8).

##### United States

| Prior | Wave 1 | Wave 2 | Wave 3 |
| --- | --- | --- | --- |
| P(*S*1\|tested) | 0.6 - 1 (0.95) | 0.5 - 1 (0.9) | 0.3 - 1 (0.8) |
| P(*S*1\|untested) | 0 - 0.15 (0.04) | 0 - 0.17 (0.055) | 0 - 0.25 (0.08) |
| *α* | 0.6 - 0.95 (0.75) | 0.65 - 1 (0.7) | 0.75 - 1 (0.9) |
| *β* | 0 - 0.4 (0.15) | 0 - 0.45 (0.15) | 0 - 0.55 (0.2) |
| P(*S*0\|test+) | 0.2 - 0.7 (0.4) | 0.25 - 0.6 (0.3) | 0.15 - 0.5 (0.25) |
| Test sensitivity (*S*e) | 0.75 - 1 (0.95) | 0.75 - 1 (0.95) | 0.7 - 1 (0.92) |
| Test specificity (*S*p) | 0.9998 - 1 (0.99995) | 0.9998 - 1 (0.99995) | 0.9998 - 1 (0.99995) |

**Table S4:** Definitions of prior distributions for the United States

###### P(S1|tested)

A case-series study of 463 patients conducted in Metropolitan Detroit during March 2020 found that 76.7% of all patients had to be hospitalized, which is indicative of the severity of their infection (Suleyman et al., 2020, p.4). As a part of the COVID-NET surveillance project in the USA which looks at hospital admissions for COVID-19 cases, a study encompassing data from 154 acute care hospitals from 13 states in March 2020 found that 45% of 2,491 hospitalized adults needed medical treatment. Overall, 86% had symptoms of cough and 80% suffered from shortness of breath (Garg et al., 2020, p.462). A study of hospital admissions for COVID-19 in the American state of Iowa from March 1 to April 4, 2020 reported that 94% had a history of fever (S. Aggarwal et al., 2020, p.92). Between March 9 and April 15, 2020, a Massachusetts community healthcare system in the United States tested 592 HCWs to discover that around 25% of all tested participants had severe symptoms of fever or shortness of breath (Lan et al., 2020, p.5). A study looking at hospital admissions for COVID-19 in California in March 2020 classified 65% of patients as urgent, emergency, or resuscitation, indicating their severity at the time of testing and admission. 91% exhibited symptoms (Tolia et al., 2020, p.505). Another study under the COVID-NET project analysing data from March-August 2020 found that around 65% of hospital admissions had symptoms of cough, chills, fever, and dyspnea (Owusu et al., 2021, p.163).

###### P(S1|untested)

A seroprevalence study conducted in the US state of Connecticut reported that 7.6-14.8% of the sample population (*N* = 567) exhibited symptoms of fever, cough, or a sore throat (Mahajan et al., 2021, p.530). A study conducted on the effects of COVID-19 on pregnant women found that 1.6% of 406,446 women hospitalized for childbirth between April-November 2020 were infected with the novel coronavirus (Jering et al., 2021, p.714).

##### α

A study involving a large community-based testing program in California conducted tests (*N*

= 547,018) on individuals self-reporting on 10 different COVID-19 symptoms from March- May 2020. 14% of participants with shortness of breath tested positive for a SARS-CoV-2 infection. Similar numbers for fever and shaking with chills were reported at 31.7% and 32.4% (Miller et al., 2021, p.15). A follow-up assessment of the same predictive symptoms conducted in December 2020 discovered that the test positivity rates for participants with more than one symptom rose to 33.9% (Miller et al., 2021, p.9). A Massachusetts community healthcare study conducted in March-April 2020 tested symptomatic HCWs and found that 10.5% of participants exhibiting severe symptoms of fever and shortness of breath tested positive for the novel coronavirus (Lan et al., 2020, p.5).

##### β

A retrospective study of healthcare-workers undergoing COVID-19 testing in Massachusetts, USA, between March-April 2020 found that 3.5% of all tested participants exhibiting mild symptoms of COVID-19 had tested positive (Lan et al., 2020, p.5). Universal screening of all pregnant women admitted at a hospital in New York (*N* = 215) in March 2020 found 13.7% of asymptomatic individuals to be SARS-CoV-2 positive (Sutton et al., 2020, p.2163). In individuals with non-severe symptoms, a Californian community-testing study observed 18.4% and 15.8% of participants exhibiting symptoms of headache and sore throat respectively to be infected with SARS-CoV-2 (Miller et al., 2021, p.15). A follow-up assessment in December of the same year discovered positivity rates for asymptomatic participants to be 8.2% (Miller et al., 2021, p.9). In June-July 2020, two different counties in the state of North Carolina, USA, conducted mass-testing of its residents (N = 68,271) to discover that 11.23% to 13.1% were infected with the novel coronavirus without any severe symptoms (Lash et al., 2020, p.1360- 1361).

###### P(S0|test +)

A Massachusetts community healthcare study conducted in March-April 2020 found that 36% of all participants with a confirmed SARS-CoV-2 infection had mild symptoms (Lan et al., 2020, p.5). Among those confirmed positive for the SARS-CoV-2 virus in a community-testing program in California conducted in March-May 2020, an average of 29% participants had non- severe symptoms (Miller et al., 2021, p.15). A symptom profile study conducted in two states in the United States in April 2020 found that 50% of the participants developed non-severe symptoms for the entire duration of illness (Yousaf et al., 2020, p.5). Lastly, CDC data from the COVID-NET network documented that 15.1% of all individuals hospitalized for COVID- 19 from March 1, 2020 to March 31, 2021 had non-severe symptoms (CDC, 2021a).

###### Test sensitivity (Se) and test specificity (Sp)

Testing in the United States was carried out using RT-PCR assays developed by the CDC. The test sensitivity and specificity priors have accordingly been informed similarly to that of Mexico defined above. Additionally, a meta-analysis of 8 studies focused on RT-PCR testing conducted in the United States using sputum, saliva, or nasopharyngeal swabs revealed a test sensitivity of 78% (Böger et al., 2021, p.25).

### Extended results

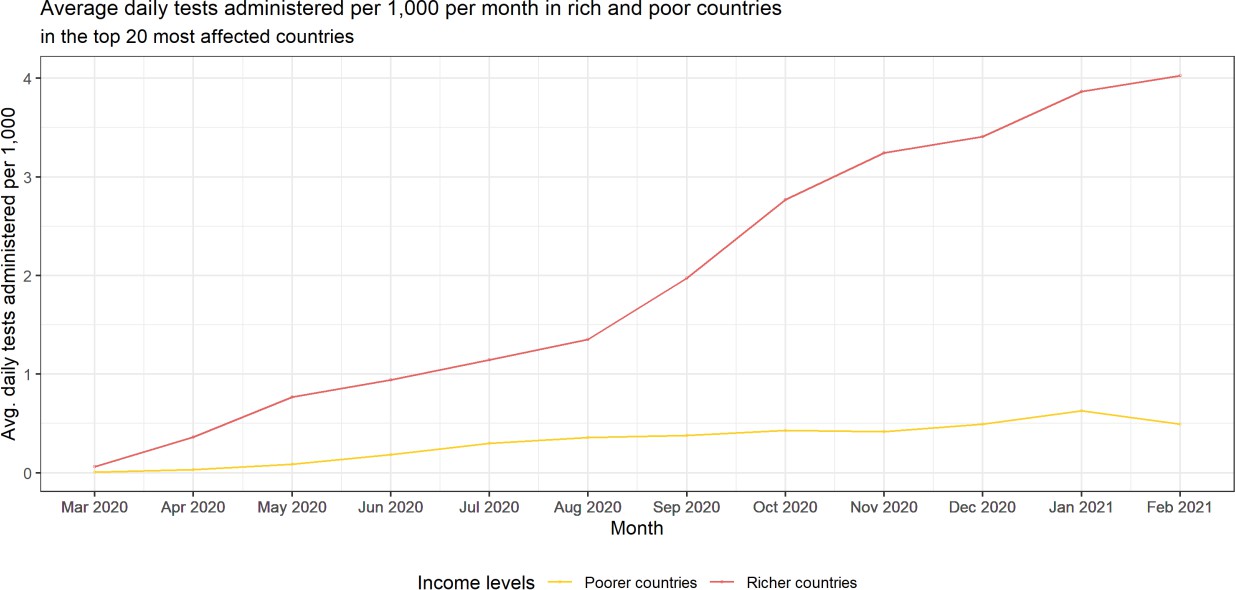
**Figure S1:** Average daily tests administered in rich and poor countries by month.

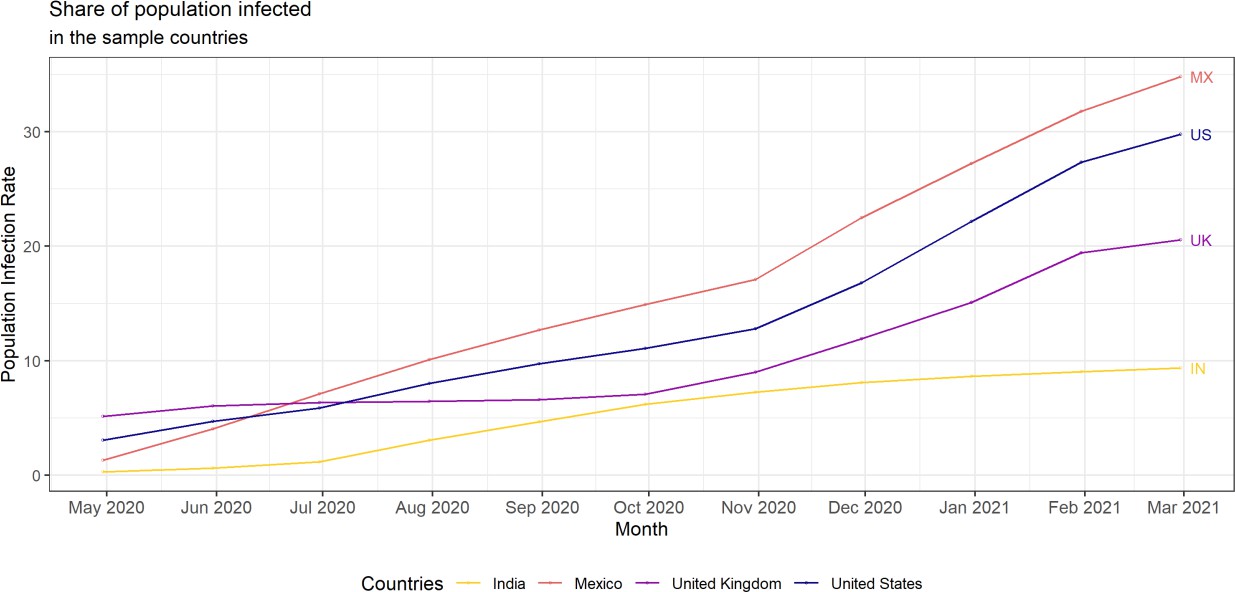
**Figure S2:** Trends in population infection rates over time.

Figure S2 presents the observed trends in the population infection rates of the four countries included in this study. We can surmise that population infection rates have been rising at a roughly constant pace since August 2020 for the countries of Mexico, the United Kingdom, and the United States. For India, we see a jump in the share of population infected from July until October 2020 after which the trend subsides. This is consistent with the surge in confirmed infections seen in the country during its first wave when the confirmed case count reached a peak of 90,000 daily cases in September 2020.

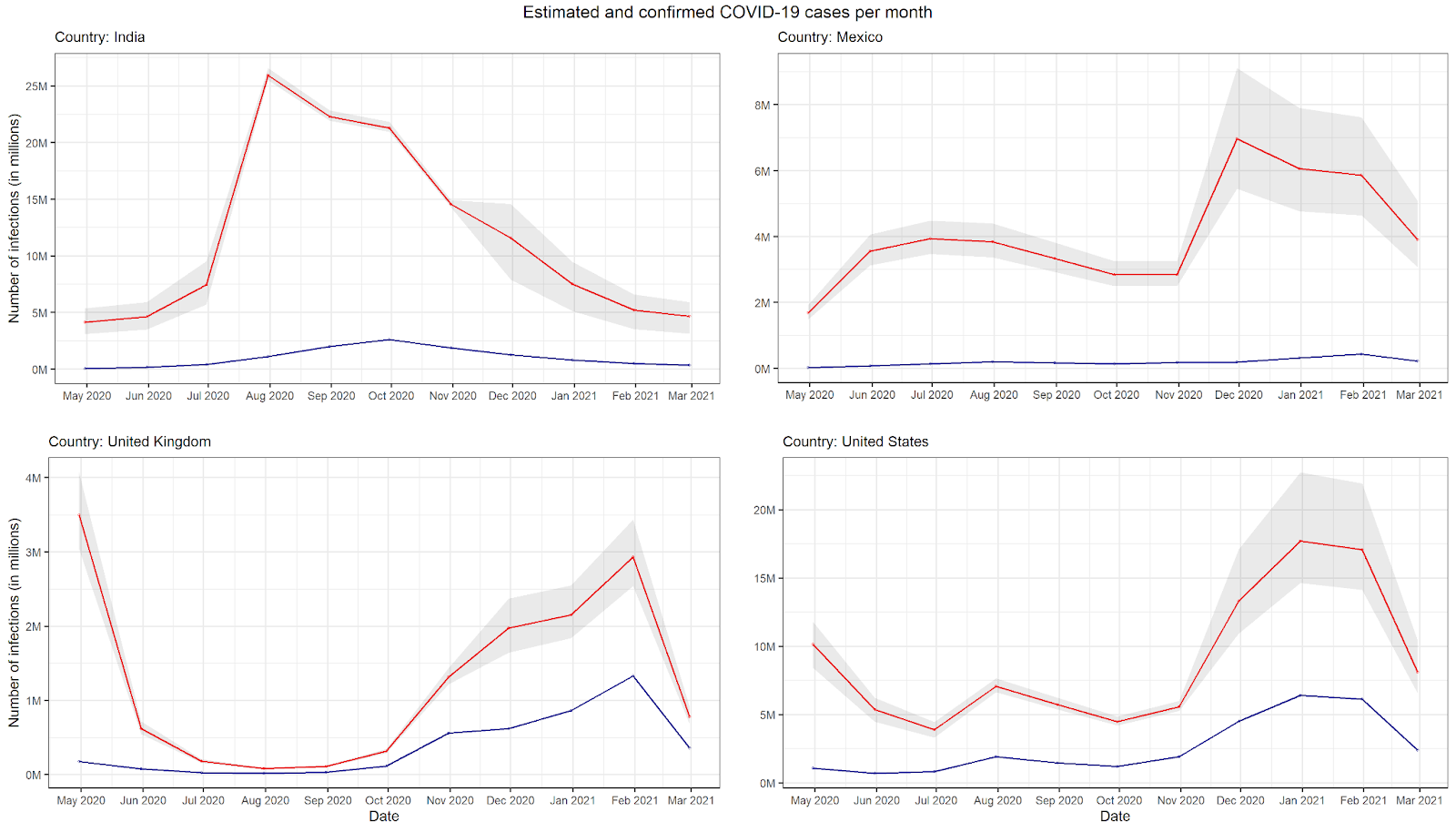

**Figure S3:** Monthly counts of estimated and confirmed cases.

**Supplementary figure 1:** Reported test-positivity rates for different countries over time.
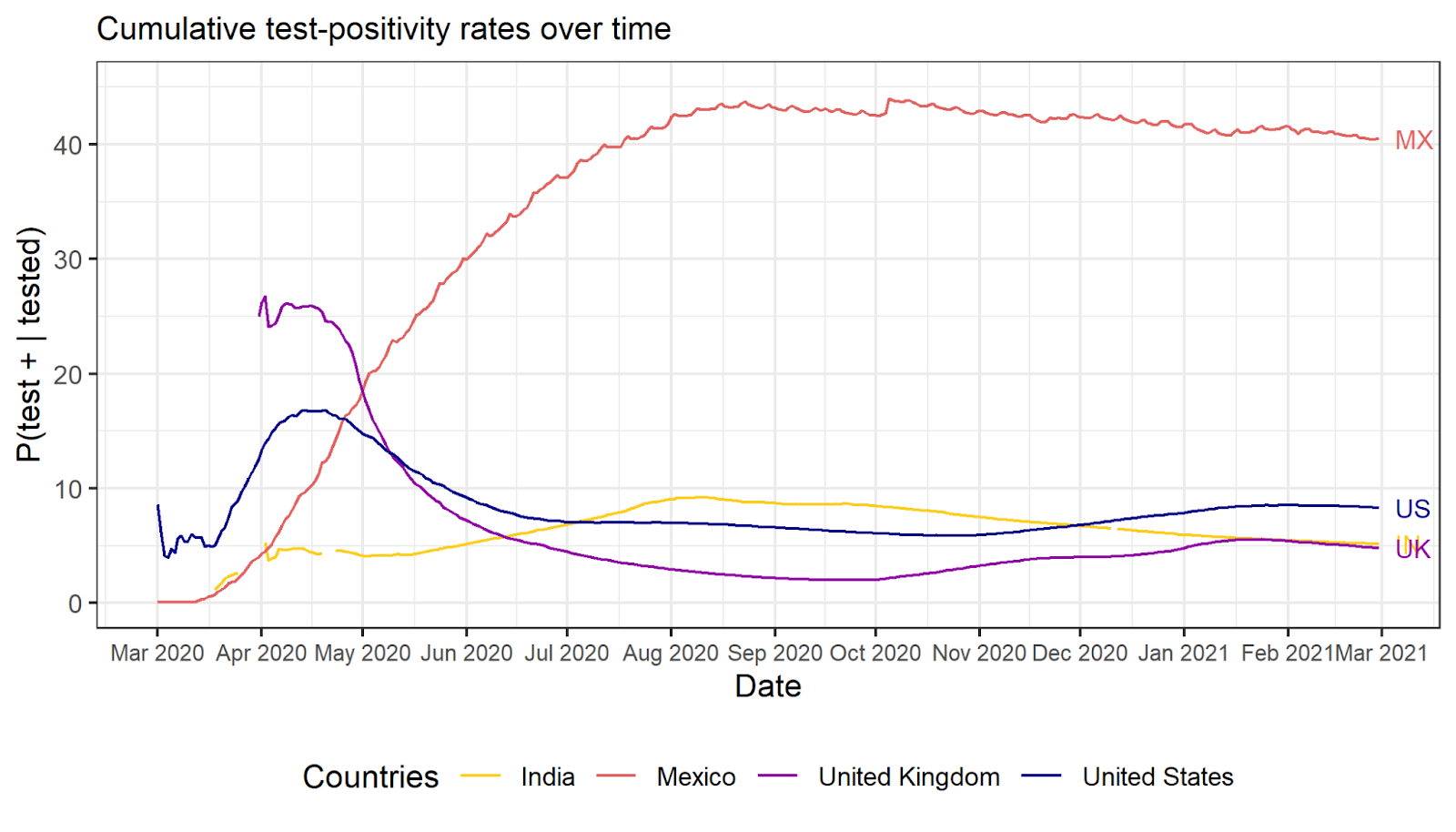

**Figure S4:** Reported test-positivity rates for different countries over time.

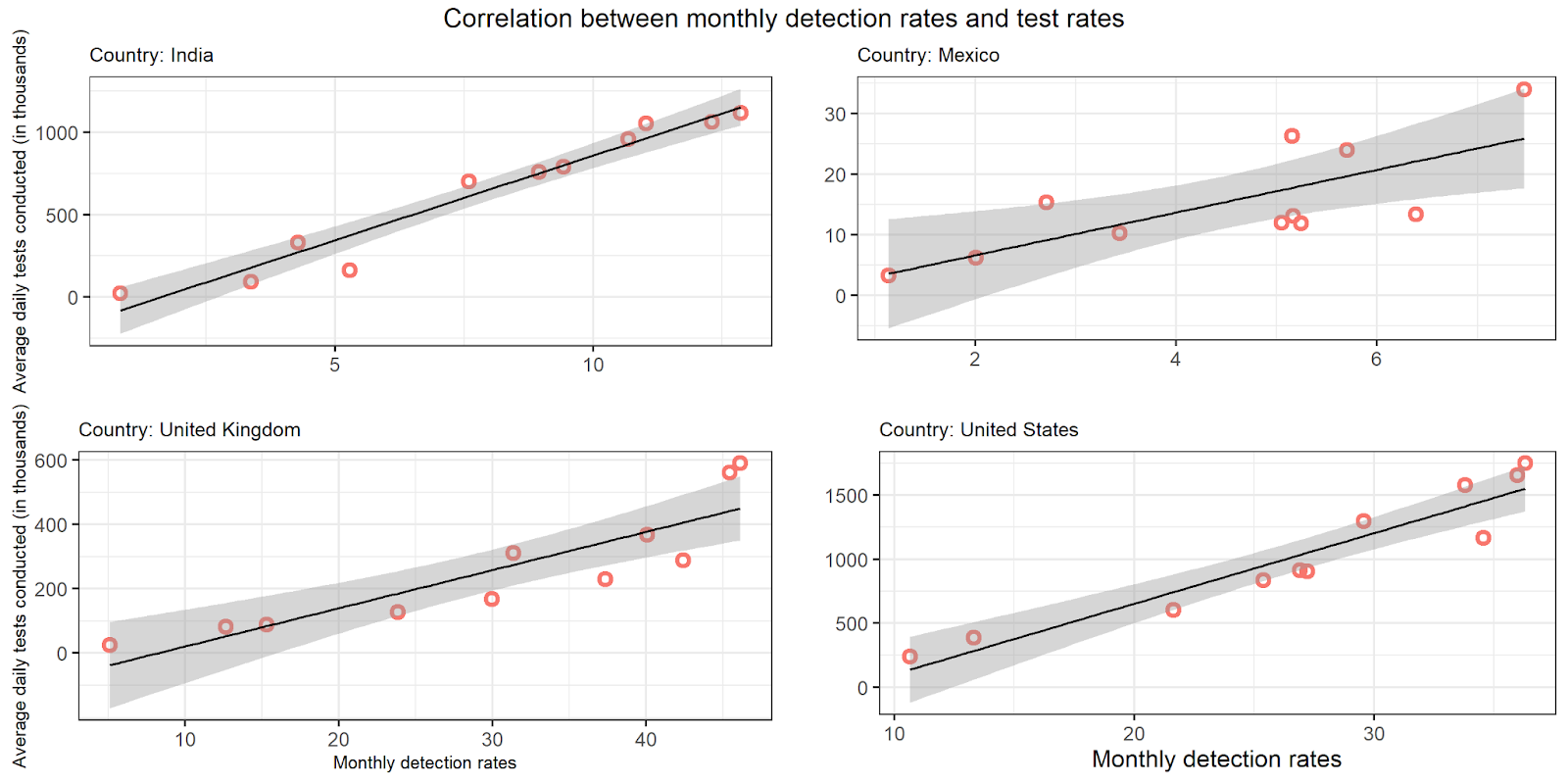

**Figure S5:** Correlation between detection rates and test positivity rates

A significant positive correlation between the monthly detection rates of true infections and the average number of daily tests conducted is found for each country; India: *r*(9) = 0.97, *p* < 0.05, Mexico: *r*(9) = 0.75, *p* < 0.05, United Kingdom: *r*(9) = 0.88, *p* < 0.05, United States: *r*(9) = 0.95, *p* < 0.05.

| **Supplementary Table 5:** Monthly cumulative estimated infections, detection rate, and population infection rate with 95% simulation interval in different countries | | | | | | |
| --- | --- | --- | --- | --- | --- | --- |
| Month | India | | | Mexico | | |
|  | Estimated infections | Detection Rate | Pop. Infection Rate | Estimated infections | Detection Rate | Pop. Infection Rate |
| Mar - April 2020 | 4,146,762 (3,093,941 -5,359,190) | 0.84 (0.65 - 1.12) | 0.30 (0.22 - 0.38) | 1,685,709 (1,482,267 - 1,925,609) | 1.14 (0.99 - 1.29) | 1.31 (1.14 - 1.49) |
| May-20 | 8,766,226 (6,580,461 -11,293,700) | 2.17 (1.69 - 2.90) | 0.64 (0.47 - 0.81) | 5,244,387 (4,615,643 - 5,986,267) | 1.73 (1.51 - 1.96) | 4.07 (3.57 - 4.64) |
| Jun-20 | 16,230,640 (12,279,586 -20,825,580) | 3.61 (2.81 - 4.76) | 1.18 (0.88 - 1.50) | 9,185,043 (8,091,297 - 10,476,140) | 2.46 (2.15 - 2.79) | 7.12 (6.27 - 8.12) |
| Jul-20 | 42,172,325 (37,805,868 -47,419,403) | 4.02 (3.57 - 4.48) | 3.06 (2.73 - 3.43) | 13,030,698 (11,461,499 - 14,880,223) | 3.26 (2.85 - 3.70) | 10.11 (8.88 - 11.50) |
| Aug-20 | 64,466,543 (59,732,531 -70,274,852) | 5.73 (5.25 - 6.17) | 4.67 (4.32 - 5.09) | 16,367,290 (14,386,095 - 18,701,225) | 3.66 (3.20 - 4.16) | 12.69 (11.11 - 14.50) |
| Sep-20 | 85,768,776 (80,678,208 - 92,114,667) | 7.36 (6.85 - 7.82) | 6.22 (5.84 - 6.67) | 19,211,827 (16,879,036 - 21,959,361) | 3.87 (3.38 - 4.40) | 14.90 (13.01 - 17.01) |
| Oct-20 | 100,318,143 (94,951,732 -107,035,848) | 8.16 (7.64 - 8.61) | 7.27 (6.88 - 7.75) | 22,056,005 (19,376,391 - 25,211,680) | 4.19 (3.66 - 4.77) | 17.11 (15.01 - 19.52) |
| Nov-20 | 111,909,198 (102,849,245 - 121,611,674) | 8.46 (7.78 - 9.20) | 8.11 (7.45 - 8.81) | 29,022,142 (24,829,447 - 34,332,784) | 3.84 (3.24 - 4.48) | 22.51 (19.22 - 26.60) |
| Dec-20 | 119,430,790 (107,974,564 - 131,092,826) | 8.60 (7.83 - 9.50) | 8.65 (7.82 - 9.49) | 35,087,155 (29,604,237 - 42,236,242) | 4.06 (3.37 - 4.81) | 27.21 (22.91 - 32.72) |
| Jan-21 | 124,639,418 (111,497,267 - 137,687,815) | 8.63 (7.81 - 9.64) | 9.03 (8.07 - 9.97) | 40,956,971 (34,250,754 - 49,858,010) | 4.55 (3.73 - 5.44) | 31.77 (26.51 - 38.63) |
| Feb-21 | 129,305,473 (114,617,974 - 143,622,760) | 8.59 (7.73 - 9.69) | 9.37 (8.30 - 10.4) | 44,863,732 (37,330,051 - 54,944,039) | 4.65 (3.79 - 5.59) | 34.80 (28.94 - 42.68) |
| Month | United Kingdom | | | United States | | |
|  | Estimated infections | Detection Rate | Pop. Infection Rate | Estimated infections | Detection Rate | Pop. Infection Rate |
| Mar - April 2020 | 3,499,026 (3,033,883 - 4,070,265) | 5.11 (4.39 - 5.89) | 5.15 (4.46 - 5.99) | 10,155,228 (8,429,436 - 11,798,885) | 10.65 (9.16 - 12.86) | 3.07 (2.54 - 3.56) |
| May-20 | 4,121,963 (3,579,785 - 4,787,052) | 6.25 (5.38 - 7.19) | 6.07 (5.27 - 7.05) | 15,546,604 (12,931,681 - 18,043,075) | 11.57 (9.96 - 13.90) | 4.70 (3.90 - 5.45) |
| Jun-20 | 4,302,847 (3,737,750 - 4,995,071) | 6.63 (5.71 - 7.63) | 6.34 (5.50 - 7.35) | 19,445,970 (16,249,095 - 22,509,299) | 13.59 (11.75 - 16.21) | 5.88 (4.90 - 6.80) |
| Jul-20 | 4,384,900 (3,808,020 - 5,087,328) | 6.95 (5.99 - 8.00) | 6.46 (5.60 - 7.49) | 26,518,013 (22,888,270 - 30,171,450) | 17.22 (15.11 - 19.94) | 8.01 (6.91 - 9.11) |
| Aug-20 | 4,496,015 (3,906,061 - 5,211,078) | 7.52 (6.48 - 8.65) | 6.62 (5.75 - 7.67) | 32,264,902 (28,277,056 - 36,403,154) | 18.68 (16.54 - 21.31) | 9.75 (8.54 - 10.90) |
| Sep-20 | 4,811,181 (4,194,601 - 5,557,422) | 9.48 (8.20 - 10.80) | 7.09 (6.17 - 8.18) | 36,756,225 (32,490,433 - 41,268,858) | 19.69 (17.50 - 22.23) | 11.11 (9.81 - 12.42) |
| Oct-20 | 6,128,495 (5,414,217 - 6,994,045) | 16.56 (14.52 - 18.77) | 9.03 (7.97 - 10.36) | 42,339,904 (37,755,733 - 47,294,601) | 21.65 (19.35 - 24.21) | 12.79 (11.41 - 14.25) |
| Nov-20 | 8,101,938 (7,056,393 - 9,364,286) | 20.17 (17.45 - 23.18) | 11.94 (10.35 - 13.70) | 55,665,966 (48,700,135 - 64,408,322) | 24.56 (21.28 - 28.47) | 16.82 (14.75 - 19.49) |
| Dec-20 | 10,254,904 (8,893,775 - 11,911,721) | 24.34 (20.94 - 28.06) | 15.11 (13.16 - 17.54) | 73,376,526 (63,351,997 - 87,133,037) | 27.39 (23.07 - 31.79) | 22.17 (19.18 - 26.34) |
| Jan-21 | 13,186,248 (11,433,609 - 15,348,937) | 29.03 (24.91 - 33.46) | 19.42 (16.82 - 22.61) | 90,463,832 (77,474,250 - 109,059,196) | 29.01 (24.06 - 33.84) | 27.33 (23.49 - 32.95) |
| Feb-21 | 13,967,911 (12,112,793 - 16,264,844) | 29.99 (25.76 - 34.52) | 20.58 (17.87 - 23.94) | 98,585,193 (84,052,809 - 119,505,261) | 29.06 (23.93 - 34.02) | 29.78 (25.36 - 36.18) |
| Notes: | Numbers for each month are representative of the total cumulative estimations as of that month. Estimations for March and April are carried out together due to a lack of sufficient data. | | | | | |

257. <https://doi.org/10.1186/s12879-021-05950-x>.

Anand, Shuchi, Maria Montez-Rath, Jialin Han, LinaCel Cadden, Patti Hunsader, Russell Kerschmann, Paul Beyer, et al. 2021. “Estimated SARS-CoV-2 Seroprevalence in US Patients Receiving Dialysis 1 Year After the Beginning of the COVID-19 Pandemic.” *JAMA Network Open* 4 (7): e2116572–e2116572. <https://doi.org/10.1001/jamanetworkopen.2021.16572>.

Anand, Shuchi, Maria Montez-Rath, Jialin Han, Julie Bozeman, Russell Kerschmann, Paul Beyer, Julie Parsonnet, and Glenn M. Chertow. 2020. “Prevalence of SARS-CoV-2 Antibodies in a Large Nationwide Sample of Patients on Dialysis in the USA: A Cross- Sectional Study.” *The Lancet* 396 (10259): 1335–44. [https://doi.org/10.1016/S0140-](https://doi.org/10.1016/S0140-6736(20)32009-2)

[6736(20)32009-2.](https://doi.org/10.1016/S0140-6736(20)32009-2)

Angulo, Frederick J., Lyn Finelli, and David L. Swerdlow. 2021. “Estimation of US SARS- CoV-2 Infections, Symptomatic Infections, Hospitalizations, and Deaths Using Seroprevalence Surveys.” *JAMA Network Open* 4 (1): e2033706. <https://doi.org/10.1001/jamanetworkopen.2020.33706>.

Antonio-Villa, Neftali Eduardo, Omar Yaxmehen Bello-Chavolla, Arsenio Vargas-Vázquez, Carlos A Fermín-Martínez, Alejandro Márquez-Salinas, and Jessica Paola Bahena-López.

2020. “Health-Care Workers with COVID-19 Living in Mexico City: Clinical Characterization and Related Outcomes.” *Clinical Infectious Diseases*, no. ciaa1487 (September). <https://doi.org/10.1093/cid/ciaa1487>.

Banerjee, Amitava, Laura Pasea, Steve Harris, Arturo Gonzalez-Izquierdo, Ana Torralbo, Laura Shallcross, Mahdad Noursadeghi, et al. 2020. “Estimating Excess 1-Year Mortality Associated with the COVID-19 Pandemic According to Underlying Conditions and Age: A Population-Based Cohort Study.” *The Lancet* 395 (10238): 1715–25. [https://doi.org/10.1016/S0140-6736(20)30854-0.](https://doi.org/10.1016/S0140-6736(20)30854-0)

Bello-Chavolla, Omar Yaxmehen, Neftali Eduardo Antonio-Villa, Arsenio Vargas-Vázquez, Carlos A Fermín-Martínez, Alejandro Márquez-Salinas, and Jessica Paola Bahena-López.

2021. “Profiling Cases With Nonrespiratory Symptoms and Asymptomatic Severe Acute Respiratory Syndrome Coronavirus 2 Infections in Mexico City.” *Clinical Infectious Diseases* 72 (10): e655–58. [https://doi.org/10.1093/cid/ciaa1288.](https://doi.org/10.1093/cid/ciaa1288)

Betsch, Cornelia, and Robert Böhm. 2016. “Detrimental Effects of Introducing Partial Compulsory Vaccination: Experimental Evidence.” *European Journal of Public Health* 26

(3): 378–81. <https://doi.org/10.1093/eurpub/ckv154>.

Bhattacharya, Sanjoy. 2010. “Reflections on the Eradication of Smallpox.” *The Lancet* 375 (9726): 1602–3. [https://doi.org/10.1016/S0140-6736(10)60692-7.](https://doi.org/10.1016/S0140-6736(10)60692-7)

Böger, Beatriz, Mariana M. Fachi, Raquel O. Vilhena, Alexandre F. Cobre, Fernanda S. Tonin, and Roberto Pontarolo. 2021. “Systematic Review with Meta-Analysis of the Accuracy of Diagnostic Tests for COVID-19.” *American Journal of Infection Control* 49 (1): 21–29. <https://doi.org/10.1016/j.ajic.2020.07.011>.

Bryant, Juliet E., Andrew S. Azman, Matthew J. Ferrari, Benjamin F. Arnold, Maciej F. Boni, Yap Boum, Kyla Hayford, et al. 2020. “Serology for SARS-CoV-2: Apprehensions, Opportunities, and the Path Forward.” *Science Immunology* 5 (47). [https://doi.org/10.1126/sciimmunol.abc6347.](https://doi.org/10.1126/sciimmunol.abc6347)

Burki, Talha Khan. 2020. “Testing for COVID-19.” *The Lancet Respiratory Medicine* 8 (7): e63–64. [https://doi.org/10.1016/S2213-2600(20)30247-2.](https://doi.org/10.1016/S2213-2600(20)30247-2)

Cardona-Pérez, J. Arturo, Isabel Villegas-Mota, A. Cecilia Helguera-Repetto, Sandra Acevedo-Gallegos, Mario Rodríguez-Bosch, Mónica Aguinaga-Ríos, Irma Coronado-Zarco, et al. 2021. “Prevalence, Clinical Features, and Outcomes of SARS-CoV-2 Infection in Pregnant Women with or without Mild/Moderate Symptoms: Results from Universal Screening in a Tertiary Care Center in Mexico City, Mexico.” *PLOS ONE* 16 (4): e0249584. [https://doi.org/10.1371/journal.pone.0249584.](https://doi.org/10.1371/journal.pone.0249584)

Centre for Disease Control and Prevention (CDC). 2021a. “COVID-19 Hospitalizations.” Accessed June 9, 2021. <https://gis.cdc.gov/grasp/COVIDNet/COVID19_5.html>.

CDC. 2021b. “CDC Washington Congressional Testimony March 4, 2020.” 2021. March 04,

2020. Accessed June 14, 2021[.](https://www.cdc.gov/washington/testimony/2020/t20200304.htm) <https://www.cdc.gov/washington/testimony/2020/t20200304.htm>.

CDC. 2020a. “Coronavirus Disease 2019 (COVID-19).” Centers for Disease Control and Prevention. February 11, 2020. Accessed June 12, 2021 <https://www.cdc.gov/coronavirus/2019-ncov/variants/variant.html>.

CDC. 2020b CDC 2019-novel coronavirus (2019-nCoV) real-time RT-PCR diagnostic panel. 2020. Accessed June 7, 2021. <https://www.fda.gov/media/134922/download>

CDC. 2020c. “Cases, Data, and Surveillance.” Centers for Disease Control and Prevention. February 11, 2020. Accessed June 14, 2021. [https://www.cdc.gov/coronavirus/2019-](https://www.cdc.gov/coronavirus/2019-ncov/covid-data/covid-net/purpose-methods.html) [ncov/covid-data/covid-net/purpose-methods.html](https://www.cdc.gov/coronavirus/2019-ncov/covid-data/covid-net/purpose-methods.html).

Cheema, Ritu, and Dean A. Blumberg. 2021. “Understanding Laboratory Testing for SARS- CoV-2.” *Children* 8 (5): 355. <https://doi.org/10.3390/children8050355>.

Chen, Albert Tian, Kevin Altschuler, Shing Hei Zhan, Yujia Alina Chan, and Benjamin E Deverman. 2021. “COVID-19 CG Enables SARS-CoV-2 Mutation and Lineage Tracking by Locations and Dates of Interest.” Edited by Jesse D Bloom and Miles P Davenport. *ELife* 10 (February): e63409. <https://doi.org/10.7554/eLife.63409>.

Cruz-Arenas, Esteban, Elizabeth Cabrera-Ruiz, Sara Laguna-Barcenas, Claudia A. Colin- Castro, Tatiana Chavez, Rafael Franco-Cendejas, Clemente Ibarra, and Javier Perez-Orive. 2021. “Serological Prevalence of SARS-CoV-2 Infection and Associated Factors in Healthcare Workers in a ‘Non-COVID’ Hospital in Mexico City.” *PLOS ONE* 16 (8): e0255916. <https://doi.org/10.1371/journal.pone.0255916>.

Dan, Jennifer M., Jose Mateus, Yu Kato, Kathryn M. Hastie, Esther Dawen Yu, Caterina E. Faliti, Alba Grifoni, et al. 2021. “Immunological Memory to SARS-CoV-2 Assessed for up to 8 Months after Infection.” *Science* 371 (6529). [https://doi.org/10.1126/science.abf4063.](https://doi.org/10.1126/science.abf4063)

Davies, Nicholas G., Christopher I. Jarvis, W. John Edmunds, Nicholas P. Jewell, Karla

Diaz-Ordaz, and Ruth H. Keogh. 2021a. “Increased Mortality in Community-Tested Cases of SARS-CoV-2 Lineage B.1.1.7.” *Nature* 593 (7858): 270–74. [https://doi.org/10.1038/s41586-](https://doi.org/10.1038/s41586-021-03426-1)

[021-03426-1.](https://doi.org/10.1038/s41586-021-03426-1)

Davies, Nicholas G., Sam Abbott, Rosanna C. Barnard, Christopher I. Jarvis, Adam J. Kucharski, James D. Munday, Carl A. B. Pearson, et al. 2021b. “Estimated Transmissibility and Impact of SARS-CoV-2 Lineage B.1.1.7 in England.” *Science* 372 (6538). [https://doi.org/10.1126/science.abg3055.](https://doi.org/10.1126/science.abg3055)

Diggle, Peter J. 2011. “Estimating Prevalence Using an Imperfect Test.” *Epidemiology Research International* 2011 (October): e608719. <https://doi.org/10.1155/2011/608719>.

Docherty, Annemarie B., Ewen M. Harrison, Christopher A. Green, Hayley E. Hardwick, Riinu Pius, Lisa Norman, Karl A. Holden, et al. 2020a. “Features of 20 133 UK Patients in Hospital with Covid-19 Using the ISARIC WHO Clinical Characterisation Protocol: Prospective Observational Cohort Study.” *BMJ* 369 (May): m1985. <https://doi.org/10.1136/bmj.m1985>.

Docherty, Annemarie B., Ewen M. Harrison, Christopher A. Green, Hayley E. Hardwick, Riinu Pius, Lisa Norman, Karl A. Holden, et al. 2020b. “Features of 16,749 Hospitalised UK Patients with COVID-19 Using the ISARIC WHO Clinical Characterisation Protocol.” *Mimeo-MedRxiv*, April, 2020.04.23.20076042. <https://doi.org/10.1101/2020.04.23.20076042>.

Fanelli, Duccio, and Francesco Piazza. 2020. “Analysis and Forecast of COVID-19 Spreading in China, Italy and France.” *Chaos, Solitons & Fractals* 134 (May): 109761. [https://doi.org/10.1016/j.chaos.2020.109761.](https://doi.org/10.1016/j.chaos.2020.109761)

Fernández-Rojas, Miguel A., Marco A. Luna-Ruiz Esparza, Abraham Campos-Romero, Diana Y. Calva-Espinosa, José L. Moreno-Camacho, Ariadna P. Langle-Martínez, Abraham García-Gil, et al. 2021. “Epidemiology of COVID-19 in Mexico: Symptomatic Profiles and Presymptomatic People.” *International Journal of Infectious Diseases* 104 (March): 572–79. <https://doi.org/10.1016/j.ijid.2020.12.086>.

Flaxman, Seth, Swapnil Mishra, Axel Gandy, H. Juliette T. Unwin, Thomas A. Mellan, Helen Coupland, Charles Whittaker, et al. 2020. “Estimating the Effects of Non-Pharmaceutical Interventions on COVID-19 in Europe.” *Nature* 584 (7820): 257–61. [https://doi.org/10.1038/s41586-020-2405-7.](https://doi.org/10.1038/s41586-020-2405-7)

Gallagher, James. 2020. “New Coronavirus Variant: What Do We Know?” *BBC News*, December 20, 2020, sec. Health. <https://www.bbc.com/news/health-55388846>.

Garg, Shikha, Lindsay Kim, Michael Whitaker, Alissa O’Halloran, Charisse Cummings, Rachel Holstein, Mila Prill, et al. 2020. “Hospitalization Rates and Characteristics of Patients Hospitalized with Laboratory-Confirmed Coronavirus Disease 2019 - COVID-NET, 14

States, March 1-30, 2020.” *MMWR. Morbidity and Mortality Weekly Report* 69 (15): 458–64. <https://doi.org/10.15585/mmwr.mm6915e3>.

Geddes, Linda, and Amanda Holpuch. 2021. “New Coronavirus Variant May Have Been in US since October.” *The Guardian*, January 1, 2021, sec. World news. [http://www.theguardian.com/world/2021/jan/01/now-coronavirus-variant-us-since-october.](http://www.theguardian.com/world/2021/jan/01/now-coronavirus-variant-us-since-october)

Gillespie, Tom, and Alix Culbertson. 2021. “Brazil COVID Variant: Where in the UK Was It Found, Is It More Deadly and Do Vaccines Work against It?” Sky News. Accessed June 8, 2021. [https://news.sky.com/story/brazil-variant-where-in-the-uk-was-it-found-is-it-more-](https://news.sky.com/story/brazil-variant-where-in-the-uk-was-it-found-is-it-more-deadly-and-do-vaccines-work-against-it-12232126) [deadly-and-do-vaccines-work-against-it-12232126.](https://news.sky.com/story/brazil-variant-where-in-the-uk-was-it-found-is-it-more-deadly-and-do-vaccines-work-against-it-12232126)

Gordon, David E., Joseph Hiatt, Mehdi Bouhaddou, Veronica V. Rezelj, Svenja Ulferts, Hannes Braberg, Alexander S. Jureka, et al. 2020. “Comparative Host-Coronavirus Protein Interaction Networks Reveal Pan-Viral Disease Mechanisms.” *Science* 370 (6521). [https://doi.org/10.1126/science.abe9403.](https://doi.org/10.1126/science.abe9403)

Gu, Youyang. 2020. “COVID-19 projections using machine learning”. [https://covid19-](https://covid19-projections.com/) [projections.com.](https://covid19-projections.com/) Accessed <26 June 2021>.

Guan, Wei-jie, Zheng-yi Ni, Yu Hu, Wen-hua Liang, Chun-quan Ou, Jian-xing He, Lei Liu, et al. 2020. “Clinical Characteristics of Coronavirus Disease 2019 in China.” *New England Journal of Medicine*, February. <https://doi.org/10.1056/NEJMoa2002032>.

Gupta, Ankesh, Surbhi Khurana, Rojaleen Das, Deepankar Srigyan, Amit Singh, Ankit Mittal, Parul Singh, et al. 2021. “Rapid Chromatographic Immunoassay-Based Evaluation of COVID-19: A Cross-Sectional, Diagnostic Test Accuracy Study & Its Implications for COVID-19 Management in India.” *Indian Journal of Medical Research* 153 (1): 126. [https://doi.org/10.4103/ijmr.IJMR_3305_20.](https://doi.org/10.4103/ijmr.IJMR_3305_20)

Gupta, Puneet, Surender Kumar, and Shashi S. Sharma. 2021. “SARS-CoV-2 Prevalence and Maternal-Perinatal Outcomes among Pregnant Women Admitted for Delivery: Experience from COVID-19-Dedicated Maternity Hospital in Jammu, Jammu and Kashmir (India).” *Journal of Medical Virology.* 2021; 1–10. <https://doi.org/10.1002/jmv.27074>.

Hansen, Christian H., Daniela Michlmayr, Sophie M. Gubbels, Kåre Mølbak, and Steen Ethelberg. 2021. “Assessment of Protection against Reinfection with SARS-CoV-2 among 4 Million PCR-Tested Individuals in Denmark in 2020: A Population-Level Observational Study.” *The Lancet* 397 (10280): 1204–12. <https://doi.org/10.1016/S0140-6736(21)00575-4>.

Hernández-Cruz, Rosa Gabriela, Daniela Sánchez-Cobo, Sandra Acevedo-Gallegos, Addy Cecilia Helguera-Repetto, Mario Roberto Rodriguez-Bosch, Victor Hugo Ramirez-Santes, Isabel Villegas-Mota, et al. 2021. “Clinical Characteristics and Risk Factors for SARS-CoV-2 Infection in Pregnant Women Attending a Third Level Reference Center in Mexico City.” *The Journal of Maternal-Fetal & Neonatal Medicine* 0 (0): 1–5. <https://doi.org/10.1080/14767058.2021.1902500>.

Huang, Chaolin, Yeming Wang, Xingwang Li, Lili Ren, Jianping Zhao, Yi Hu, Li Zhang, et al. 2020. “Clinical Features of Patients Infected with 2019 Novel Coronavirus in Wuhan, China.” *The Lancet* 395 (10223): 497–506. <https://doi.org/10.1016/S0140-6736(20)30183-5>.

Indian Council of Medical Research (ICMR). 2020. “Guidance for appropriate recording of COVID-19 related deaths in India”. Indian Council of Medical Research. 2020. Accessed July 3, 2021. <https://ncdirindia.org/Downloads/CoD_COVID-19_Guidance.pdf>

Institute for Health Metrics and Evaluation (IHME). 2021. “Estimation of Excess Mortality Due to COVID-19.” 2021. Institute for Health Metrics and Evaluation. April 22, 2021. [http://www.healthdata.org/special-analysis/estimation-excess-mortality-due-covid-19-and-](http://www.healthdata.org/special-analysis/estimation-excess-mortality-due-covid-19-and-scalars-reported-covid-19-deaths) [scalars-reported-covid-19-deaths.](http://www.healthdata.org/special-analysis/estimation-excess-mortality-due-covid-19-and-scalars-reported-covid-19-deaths)

Jering, Karola S., Brian L. Claggett, Jonathan W. Cunningham, Ning Rosenthal, Orly Vardeny, Michael F. Greene, and Scott D. Solomon. 2021. “Clinical Characteristics and Outcomes of Hospitalized Women Giving Birth With and Without COVID-19.” *JAMA Internal Medicine* 181 (5): 714–17. <https://doi.org/10.1001/jamainternmed.2020.9241>.

Jha, Sujeet, Aakriti Soni, Samreen Siddiqui, Nitish Batra, Nikita Goel, Sneha Dey, Sandeep Budhiraja, and Rahul Naithani. 2020. “Prevalence of Flu-like Symptoms and COVID-19 in

Healthcare Workers from India.” *The Journal of the Association of Physicians of India* 68

(7): 27–29.

Johansson, Michael A., Talia M. Quandelacy, Sarah Kada, Pragati Venkata Prasad, Molly Steele, John T. Brooks, Rachel B. Slayton, Matthew Biggerstaff, and Jay C. Butler. 2021. “SARS-CoV-2 Transmission From People Without COVID-19 Symptoms.” *JAMA Network Open* 4 (1): e2035057. <https://doi.org/10.1001/jamanetworkopen.2020.35057>.

Joshi, Ashish, Apeksha H. Mewani, Srishti Arora, and Ashoo Grover. 2021. “India’s COVID-19 Burdens, 2020.” *Frontiers in Public Health* 9: 294. <https://doi.org/10.3389/fpubh.2021.608810>.

Katz, Josh, Denise Lu, and Margot Sanger-Katz. 2020. “U.S. Coronavirus Death Toll Is Far Higher Than Reported, C.D.C. Data Suggests.” *The New York Times*, April 28, 2020, sec.

U.S. <https://www.nytimes.com/interactive/2020/04/28/us/coronavirus-death-toll-total.html>.

Kayina, Choro Athiphro, Damarla Haritha, Lipika Soni, Srikant Behera, Parvathy Ramachandran Nair, M. Gouri, Kavitha Girish, et al. 2020. “Epidemiological & Clinical Characteristics & Early Outcome of COVID-19 Patients in a Tertiary Care Teaching Hospital in India: A Preliminary Analysis.” *Indian Journal of Medical Research* 152 (1): 100. [https://doi.org/10.4103/ijmr.IJMR_2890_20.](https://doi.org/10.4103/ijmr.IJMR_2890_20)

Keeley, Alexander J., Cariad Evans, Hayley Colton, Michael Ankcorn, Alison Cope, Amy State, Tracy Bennett, Prosenjit Giri, Thushan I. de Silva, and Mohammad Raza. 2020. “Roll- out of SARS-CoV-2 Testing for Healthcare Workers at a Large NHS Foundation Trust in the United Kingdom, March 2020.” *Eurosurveillance* 25 (14): 2000433. <https://doi.org/10.2807/1560-7917.ES.2020.25.14.2000433>.

Khalil, Asma, Robert Hill, Shamez Ladhani, Katherine Pattisson, and Pat O’Brien. 2021. “COVID-19 Screening of Health-Care Workers in a London Maternity Hospital.” *The Lancet Infectious Diseases* 21 (1): 23–24. <https://doi.org/10.1016/S1473-3099(20)30403-5>.

Krishnasamy, Narayanasamy, Murugan Natarajan, Arunkumar Ramachandran, Jeromie Wesley Vivian Thangaraj, Theranirajan Etherajan, Jayanthi Rengarajan, Meenakshi

Shanmugasundaram, et al. 2020. “Clinical Outcomes among Asymptomatic or Mildly Symptomatic COVID-19 Patients in an Isolation Facility in Chennai, India.” *The American Journal of Tropical Medicine and Hygiene* 104 (1): 85–90. [https://doi.org/10.4269/ajtmh.20-](https://doi.org/10.4269/ajtmh.20-1096) [1096](https://doi.org/10.4269/ajtmh.20-1096).

Kumar, G. Anil, Lalit Dandona, and Rakhi Dandona. 2019. “Completeness of Death Registration in the Civil Registration System, India (2005 to 2015).” *The Indian Journal of Medical Research* 149 (6): 740–47. <https://doi.org/10.4103/ijmr.IJMR_1620_17>.

Lan, Fan-Yun, Robert Filler, Soni Mathew, Jane Buley, Eirini Iliaki, Lou Ann Bruno-Murtha, Rebecca Osgood, Costas A. Christophi, Alejandro Fernandez-Montero, and Stefanos N. Kales. 2020. “COVID-19 Symptoms Predictive of Healthcare Workers’ SARS-CoV-2 PCR Results.” *PLOS ONE* 15 (6): e0235460. <https://doi.org/10.1371/journal.pone.0235460>.

Lash, R. Ryan, Catherine V. Donovan, Aaron T. Fleischauer, Zack S. Moore, Gibbie Harris, Susan Hayes, Meg Sullivan, et al. 2020. “COVID-19 Contact Tracing in Two Counties — North Carolina, June–July 2020.” MMWR. Morbidity and Mortality Weekly Report 69 (38): 1360–63. <https://doi.org/10.15585/mmwr.mm6938e3>.

Lash, Timothy L., Matthew P. Fox, and Aliza K. Fink, 2009. “Applying Quantitative Bias Analysis to Epidemiologic Data”. Dordrecht: Springer.

Linka, Kevin, Mathias Peirlinck, and Ellen Kuhl. 2020. “The Reproduction Number of COVID-19 and Its Correlation with Public Health Interventions.” *Computational Mechanics* 66 (4): 1035–50. [https://doi.org/10.1007/s00466-020-01880-8.](https://doi.org/10.1007/s00466-020-01880-8)

Levin, Andrew T., William P. Hanage, Nana Owusu-Boaitey, Kensington B. Cochran, Seamus P. Walsh, and Gideon Meyerowitz-Katz. 2020. “Assessing the Age Specificity of Infection Fatality Rates for COVID-19: Systematic Review, Meta-Analysis, and Public

Policy Implications.” *European Journal of Epidemiology* 35 (12): 1123–38. [https://doi.org/10.1007/s10654-020-00698-1.](https://doi.org/10.1007/s10654-020-00698-1)

Lewis, Megan, Ruth Sanchez, Sarah Auerbach, Dolly Nam, Brennan Lanier, Jeffrey Taylor, Cynthia Jaso, et al. 2020. “COVID-19 Outbreak Among College Students After a Spring Break Trip to Mexico — Austin, Texas, March 26–April 5, 2020.” *MMWR. Morbidity and Mortality Weekly Report* 69. <https://doi.org/10.15585/mmwr.mm6926e1>.

MacLehose, Richard F., and Paul Gustafson. 2012. “Is Probabilistic Bias Analysis Approximately Bayesian?” *Epidemiology* 23 (1): 151–58. [https://doi.org/10.1097/EDE.0b013e31823b539c.](https://doi.org/10.1097/EDE.0b013e31823b539c)

Mahajan, Shiwani, Rajesh Srinivasan, Carrie A. Redlich, Sara K. Huston, Kelly M. Anastasio, Lisa Cashman, Dorothy S. Massey, et al. 2021. “Seroprevalence of SARS-CoV-2- Specific IgG Antibodies Among Adults Living in Connecticut: Post-Infection Prevalence (PIP) Study.” *The American Journal of Medicine* 134 (4): 526-534.e11. <https://doi.org/10.1016/j.amjmed.2020.09.024>.

Mahajan, Niraj, Rahul Gajbhiye, Pradip Lokhande, Shubhada Bahirat, Kshitija Mahajan, Shailesh Mohite, Surbhi Rathi, Vishal Rakh, Gauri Patokar, and Arundhati Tilve. 2020. “Prevalence and Clinical Presentation of COVID-19 among Healthcare Workers at a Dedicated Hospital in India.” *The Journal of the Association of Physicians of India* 68 (December): 16–21.

Malani, Anup, Daksha Shah, Gagandeep Kang, Gayatri Nair Lobo, Jayanthi Shastri, Manoj Mohanan, Rajesh Jain, et al. 2020. “Seroprevalence of SARS-CoV-2 in Slums and Non- Slums of Mumbai, India, during June 29-July 19, 2020.” *Mimeo-MedRxiv*, September, 2020.08.27.20182741. <https://doi.org/10.1101/2020.08.27.20182741>.

Martinez-Fierro, Margarita L., Jorge Ríos-Jasso, Idalia Garza-Veloz, Lucia Reyes-Veyna, Rosa Maria Cerda-Luna, Iliana Duque-Jara, Maribel Galvan-Jimenez, et al. 2021a. “The Role of Close Contacts of COVID-19 Patients in the SARS-CoV-2 Transmission: An Emphasis on the Percentage of Nonevaluated Positivity in Mexico.” *American Journal of Infection Control* 49 (1): 15–20. <https://doi.org/10.1016/j.ajic.2020.10.002>.

Martinez-Fierro, Margarita L., Martha Diaz-Lozano, Claudia Alvarez-Zuñiga, Leticia A. Ramirez-Hernandez, Roxana Araujo-Espino, Perla M. Trejo-Ortiz, Fabiana E. Mollinedo- Montaño, et al. 2021b. “Population-Based COVID-19 Screening in Mexico: Assessment of Symptoms and Their Weighting in Predicting SARS-CoV-2 Infection.” *Medicina* 57 (4): 363. [https://doi.org/10.3390/medicina57040363.](https://doi.org/10.3390/medicina57040363)

McEvoy, Jemima. 2020. “WHO Estimates Coronavirus Infected 10% Of World’s Population.” Forbes. Accessed July 19, 2021. [https://www.forbes.com/sites/jemimamcevoy/2020/10/05/who-estimates-coronavirus-](https://www.forbes.com/sites/jemimamcevoy/2020/10/05/who-estimates-coronavirus-infected-10-of-worlds-population/) [infected-10-of-worlds-population/](https://www.forbes.com/sites/jemimamcevoy/2020/10/05/who-estimates-coronavirus-infected-10-of-worlds-population/).

Menni, Cristina, Ana M. Valdes, Maxim B. Freidin, Carole H. Sudre, Long H. Nguyen, David A. Drew, Sajaysurya Ganesh, et al. 2020. “Real-Time Tracking of Self-Reported Symptoms to Predict Potential COVID-19.” *Nature Medicine* 26 (7): 1037–40. [https://doi.org/10.1038/s41591-020-0916-2.](https://doi.org/10.1038/s41591-020-0916-2)

Meril Diagnostics. 2020. “Meril COVID-19 One-step RT-PCR Kit”. Accessed June 15, 2021. <https://www.merillife.com/assets/pdfs/clinical-data/pack-insert-iud-1608117820pdf.pdf>

Meyerowitz-Katz, Gideon, and Lea Merone. 2020. “A Systematic Review and Meta-Analysis of Published Research Data on COVID-19 Infection Fatality Rates.” *International Journal of Infectious Diseases* 101 (December): 138–48. <https://doi.org/10.1016/j.ijid.2020.09.1464>.

Miller, Dave P., Scott Morrow, Robert M. Califf, Cameron Kaiser, Ritu Kapur, Casimir Starsiak, Jessica Mega, and William J. Marks. 2021. “COVID-19 Test Positivity: Predictive Value of Various Symptoms in a Large Community-Based Testing Program in California.” *Mimeo-MedRxiv*, March, 2021.03.03.21252014. <https://doi.org/10.1101/2021.03.03.21252014>.

Miller, Tyler E., Wilfredo F. Garcia Beltran, Adam Z. Bard, Tasos Gogakos, Melis N. Anahtar, Michael Gerino Astudillo, Diane Yang, et al. 2020. “Clinical Sensitivity and Interpretation of PCR and Serological COVID-19 Diagnostics for Patients Presenting to the Hospital.” *The FASEB Journal* 34 (10): 13877–84. <https://doi.org/10.1096/fj.202001700RR>.

Office for National Statistics. 2021a. “Coronavirus (COVID-19) Infections in the Community in the UK - Office for National Statistics.” Accessed June 14, 2021. [https://www.ons.gov.uk/peoplepopulationandcommunity/healthandsocialcare/conditionsandd](https://www.ons.gov.uk/peoplepopulationandcommunity/healthandsocialcare/conditionsanddiseases/datasets/coronaviruscovid19infectionsinthecommunityinengland) [iseases/datasets/coronaviruscovid19infectionsinthecommunityinengland](https://www.ons.gov.uk/peoplepopulationandcommunity/healthandsocialcare/conditionsanddiseases/datasets/coronaviruscovid19infectionsinthecommunityinengland).

Office for National Statistics. 2021b. “Coronavirus (COVID-19) Infection Survey, antibody data for the UK: 3 February 2021.” Accessed August 21, 2021. [https://www.ons.gov.uk/peoplepopulationandcommunity/healthandsocialcare/conditionsandd](https://www.ons.gov.uk/peoplepopulationandcommunity/healthandsocialcare/conditionsanddiseases/articles/coronaviruscovid19infectionsurveyantibodydatafortheuk/3february2021) [iseases/articles/coronaviruscovid19infectionsurveyantibodydatafortheuk/3february2021](https://www.ons.gov.uk/peoplepopulationandcommunity/healthandsocialcare/conditionsanddiseases/articles/coronaviruscovid19infectionsurveyantibodydatafortheuk/3february2021)

Office for National Statistics. 2021c. “COVID-19 Infection Survey: methods and further information.” Accessed August 18, 2021. [https://www.ons.gov.uk/peoplepopulationandcommunity/healthandsocialcare/conditionsandd](https://www.ons.gov.uk/peoplepopulationandcommunity/healthandsocialcare/conditionsanddiseases/methodologies/covid19infectionsurveypilotmethodsandfurtherinformation#test-sensitivity-and-specificity) [iseases/methodologies/covid19infectionsurveypilotmethodsandfurtherinformation#test-](https://www.ons.gov.uk/peoplepopulationandcommunity/healthandsocialcare/conditionsanddiseases/methodologies/covid19infectionsurveypilotmethodsandfurtherinformation#test-sensitivity-and-specificity) [sensitivity-and-specificity](https://www.ons.gov.uk/peoplepopulationandcommunity/healthandsocialcare/conditionsanddiseases/methodologies/covid19infectionsurveypilotmethodsandfurtherinformation#test-sensitivity-and-specificity)

Ortiz-Brizuela, Edgar, Marco Villanueva-Reza, María F. González-Lara, Karla M. Tamez- Torres, Carla M. Román-Montes, Bruno A. Díaz-Mejía, Esteban Pérez-García, et al. 2020.

“Clinical and Epidemiological Characteristics of Patients Diagnosed with COVID-19 in a Tertiary Care Center in Mexico City: A Prospective Cohort Study.” *Revista de Investigación Clínica* 72 (3): 165–77. <https://doi.org/10.24875/ric.20000211>.

O’Toole, Áine, Verity Hill, Oliver G. Pybus, Alexander Watts, Issac I. Bogoch, Kamran Khan, Jane P. Messina, et al. 2021. “Tracking the International Spread of SARS-CoV-2 Lineages B.1.1.7 and B.1.351/501Y-V2.” Accessed June 12 2021. [https://cov-](https://cov-lineages.org/global_report_B.1.1.7.html) [lineages.org/global_report_B.1.1.7.html](https://cov-lineages.org/global_report_B.1.1.7.html), <https://cov-lineages.org/global_report_B.1.351.html>, and <https://cov-lineages.org/global_report_P.1.html>

Owusu, Daniel, Lindsay Kim, Alissa O’Halloran, Michael Whitaker, Alexandra M Piasecki, Arthur Reingold, Nisha B Alden, et al. 2021. “Characteristics of Adults Aged 18–49 Years Without Underlying Conditions Hospitalized With Laboratory-Confirmed Coronavirus Disease 2019 in the United States: COVID-NET—March–August 2020.” *Clinical Infectious Diseases* 72 (5): e162–66. [https://doi.org/10.1093/cid/ciaa1806.](https://doi.org/10.1093/cid/ciaa1806)

Peto, Julian, James Carpenter, George Davey Smith, Stephen Duffy, Richard Houlston, David J. Hunter, Klim McPherson, et al. 2020. “Weekly COVID-19 Testing with Household Quarantine and Contact Tracing Is Feasible and Would Probably End the Epidemic.” *Royal Society Open Science* 7 (6): 200915. <https://doi.org/10.1098/rsos.200915>.

Phipps, Steven J., R. Quentin Grafton, and Tom Kompas. 2020. “Robust Estimates of the True (Population) Infection Rate for COVID-19: A Backcasting Approach.” *Royal Society Open Science* 7 (11): 200909. <https://doi.org/10.1098/rsos.200909>.

Poole, David, and Adrian E. Raftery. 2000. “Inference for Deterministic Simulation Models: The Bayesian Melding Approach.” *Journal of the American Statistical Association* 95 (452): 1244–55. <https://doi.org/10.1080/01621459.2000.10474324>.

Pouwels, Koen B., Thomas House, Emma Pritchard, Julie V. Robotham, Paul J. Birrell, Andrew Gelman, Karina-Doris Vihta, et al. 2021. “Community Prevalence of SARS-CoV-2 in England from April to November, 2020: Results from the ONS Coronavirus Infection Survey.” *The Lancet Public Health* 6 (1): e30–38. [https://doi.org/10.1016/S2468-](https://doi.org/10.1016/S2468-2667(20)30282-6) [2667(20)30282-6.](https://doi.org/10.1016/S2468-2667(20)30282-6)

Rambaut, Andrew, Edward C. Holmes, Áine O’Toole, Verity Hill, John T. McCrone, Christopher Ruis, Louis du Plessis, and Oliver G. Pybus. 2020. “A Dynamic Nomenclature Proposal for SARS-CoV-2 Lineages to Assist Genomic Epidemiology.” *Nature Microbiology* 5 (11): 1403–7. [https://doi.org/10.1038/s41564-020-0770-5.](https://doi.org/10.1038/s41564-020-0770-5)

Reese, Heather, A Danielle Iuliano, Neha N Patel, Shikha Garg, Lindsay Kim, Benjamin J Silk, Aron J Hall, Alicia Fry, and Carrie Reed. 2021. “Estimated Incidence of Coronavirus Disease 2019 (COVID-19) Illness and Hospitalization—United States, February–September 2020.” *Clinical Infectious Diseases* 72 (12): e1010–17. <https://doi.org/10.1093/cid/ciaa1780>.

Reiner, Robert C., Ryan M. Barber, James K. Collins, Peng Zheng, Christopher Adolph, James Albright, Catherine M. Antony, et al. 2021. “Modeling COVID-19 Scenarios for the United States.” *Nature Medicine* 27 (1): 94–105. [https://doi.org/10.1038/s41591-020-1132-9.](https://doi.org/10.1038/s41591-020-1132-9)

Riley, Steven, Kylie E. C. Ainslie, Oliver Eales, Benjamin Jeffrey, Caroline E. Walters, Christina Atchison, Peter J. Diggle, et al. 2020. “Community Prevalence of SARS-CoV-2 Virus in England during May 2020: REACT Study.” *Mimeo-MedRxiv*, July, 2020.07.10.20150524. <https://doi.org/10.1101/2020.07.10.20150524>.

Ritchie, Hannah, Esteban Ortiz-Ospina, Diana Beltekian, Edouard Mathieu, Joe Hasell, Bobbie Macdonald, Charlie Giattino, Cameron Appel, Lucas Rodés-Guirao and Max Roser. 2020. "Coronavirus Pandemic (COVID-19)". *Published online at OurWorldInData.org.*

Accessed July 3, 2021. Retrieved from: ['https://ourworldindata.org/coronavirus](https://ourworldindata.org/coronavirus)’ [Online Resource]

Rodda, Lauren B., Jason Netland, Laila Shehata, Kurt B. Pruner, Peter A. Morawski, Christopher D. Thouvenel, Kennidy K. Takehara, et al. 2021. “Functional SARS-CoV-2- Specific Immune Memory Persists after Mild COVID-19.” *Cell* 184 (1): 169-183.e17. [https://doi.org/10.1016/j.cell.2020.11.029.](https://doi.org/10.1016/j.cell.2020.11.029)

Romer, Paul. 2020. “Roadmap to Responsibly Reopen America.” 23 April 2020. Accessed July 2, 2021. <https://paulromer.net/roadmap-to-reopen-america/>.

Romero-Gameros, Carlos Alfonso, Salomón Waizel-Haiat, Victoria Mendoza-Zubieta, Alfredo Anaya-Dyck, Mayra Alejandra López-Moreno, Tania Colin-Martinez, José Luis Martínez-Ordaz, et al. 2020. “Evaluation of Predictive Value of Olfactory Dysfunction, as a Screening Tool for COVID-19.” *Laryngoscope Investigative Otolaryngology* 5 (6): 983–91. <https://doi.org/10.1002/lio2.482>.

Rothman, Kenneth J., Sander Greenland, and Timothy L. Lash, *Modern Epidemiology*, Lippincott Williams & Wilkins, Philadelphia, Pa, USA, 3rd edition, 2008.

Russell, Timothy W., Nick Golding, Joel Hellewell, Sam Abbott, Lawrence Wright, Carl A.

B. Pearson, Kevin van Zandvoort, et al. 2020. “Reconstructing the Early Global Dynamics of Under-Ascertained COVID-19 Cases and Infections.” *BMC Medicine* 18 (1): 332. [https://doi.org/10.1186/s12916-020-01790-9.](https://doi.org/10.1186/s12916-020-01790-9)

S, Rukmini. 2021. “We Still Don’t Know India’s True COVID-19 Death Toll.” *Foreign Policy* (blog). Accessed July 3, 2021. [https://foreignpolicy.com/2021/06/30/india-](https://foreignpolicy.com/2021/06/30/india-coronavirus-pandemic-case-fatality-rate-data-undercounting-modi-vardhan-bjp/) [coronavirus-pandemic-case-fatality-rate-data-undercounting-modi-vardhan-bjp/](https://foreignpolicy.com/2021/06/30/india-coronavirus-pandemic-case-fatality-rate-data-undercounting-modi-vardhan-bjp/).

Sample, Ian, Natalie Grover. 23 December 2020. "South African Covid-19 variant has reached the UK, says Matt Hancock". *The Guardian*. London. Accessed June 8, 2021. [https://www.theguardian.com/world/2020/dec/23/south-african-covid-19-variant-has-](https://www.theguardian.com/world/2020/dec/23/south-african-covid-19-variant-has-reached-the-uk-says-matt-hancock) [reached-the-uk-says-matt-hancock](https://www.theguardian.com/world/2020/dec/23/south-african-covid-19-variant-has-reached-the-uk-says-matt-hancock)

Saxena, Sonal, Vikas Manchanda, Tanu Sagar, Nazia Nagi, Oves Siddiqui, Abhishek Yadav, Nitin Arora, et al. 2021. “Clinical Characteristic and Epidemiological Features of SARS CoV-2 Disease Patients from a COVID-19 Designated Hospital in New Delhi.” *Journal of Medical Virology* 93 (4): 2487–92. <https://doi.org/10.1002/jmv.26777>.

Sharma, Arvind Kumar, Asrar Ahmed, Vaseem Naheed Baig, Prahalad Dhakad, Gaurav Dalela, Sudhanshu Kacker, Vasim Raja Panwar, Raja Babu Panwar, and Rajeev Gupta. 2020. “Characteristics and Outcomes of Hospitalized Young Adults with Mild Covid-19.” *Mimeo- MedRxiv*, June, 2020.06.02.20106310. <https://doi.org/10.1101/2020.06.02.20106310>.

Smith, Louise E., Henry W. W. Potts, Richard Amlôt, Nicola T. Fear, Susan Michie, and G. James Rubin. 2021. “Adherence to the Test, Trace, and Isolate System in the UK: Results

from 37 Nationally Representative Surveys.” *BMJ* 372 (March): n608. <https://doi.org/10.1136/bmj.n608>.

Soni, Shiv Lal, Kamal Kajal, L. N. Yaddanapudi, Pankaj Malhotra, Goverdhan Dutt Puri, Ashish Bhalla, Mini P. Singh, et al. 2021. “Demographic & Clinical Profile of Patients with COVID-19 at a Tertiary Care Hospital in North India.” *Indian Journal of Medical Research* 153 (1): 115. <https://doi.org/10.4103/ijmr.IJMR_2311_20>.

Steenhuysen, Julie. 2021. “U.S. Data Show Rising ‘breakthrough’ Infections among Fully Vaccinated.” *Reuters*, August 25, 2021, sec. Healthcare & Pharmaceuticals. [https://www.reuters.com/business/healthcare-pharmaceuticals/us-data-show-rising-](https://www.reuters.com/business/healthcare-pharmaceuticals/us-data-show-rising-breakthrough-infections-among-fully-vaccinated-2021-08-24/) [breakthrough-infections-among-fully-vaccinated-2021-08-24/](https://www.reuters.com/business/healthcare-pharmaceuticals/us-data-show-rising-breakthrough-infections-among-fully-vaccinated-2021-08-24/).

Suleyman, Geehan, Raef A. Fadel, Kelly M. Malette, Charles Hammond, Hafsa Abdulla, Abigail Entz, Zachary Demertzis, et al. 2020. “Clinical Characteristics and Morbidity Associated With Coronavirus Disease 2019 in a Series of Patients in Metropolitan Detroit.” *JAMA Network Open* 3 (6). <https://doi.org/10.1001/jamanetworkopen.2020.12270>.

Sutton, Desmond, Karin Fuchs, Mary D’Alton, and Dena Goffman. 2020. “Universal Screening for SARS-CoV-2 in Women Admitted for Delivery.” *New England Journal of Medicine*, April. <https://doi.org/10.1056/NEJMc2009316>.

Tahan, Stephen, Bijal A. Parikh, Lindsay Droit, Meghan A. Wallace, Carey-Ann D. Burnham, and David Wang. 2021 “SARS-CoV-2 E Gene Variant Alters Analytical Sensitivity Characteristics of Viral Detection Using a Commercial Reverse Transcription- PCR Assay.” *Journal of Clinical Microbiology* 59 (7): e00075-21. <https://doi.org/10.1128/JCM.00075-21>.

Thorne, Lucy G., Mehdi Bouhaddou, Ann-Kathrin Reuschl, Lorena Zuliani-Alvarez, Benjamin J. Polacco, Adrian Pelin, Jyoti Batra, et al. 2021. “Evolution of Enhanced Innate Immune Evasion by the SARS-CoV-2 B.1.1.7 UK Variant.” *Mimeo-BioRxiv*, June, 2021.06.06.446826. <https://doi.org/10.1101/2021.06.06.446826>.

Tolia, Vaishal M., Theodore C. Chan, and Edward M. Castillo. 2020. “Preliminary Results of Initial Testing for Coronavirus (COVID-19) in the Emergency Department.” *The Western Journal of Emergency Medicine* 21 (3): 503–6. <https://doi.org/10.5811/westjem.2020.3.47348>.

Treibel, Thomas A., Charlotte Manisty, Maudrian Burton, Áine McKnight, Jonathan Lambourne, João B. Augusto, Xosé Couto-Parada, Teresa Cutino-Moguel, Mahdad Noursadeghi, and James C. Moon. 2020. “COVID-19: PCR Screening of Asymptomatic Health-Care Workers at London Hospital.” *The Lancet* 395 (10237): 1608–10. [https://doi.org/10.1016/S0140-6736(20)31100-4.](https://doi.org/10.1016/S0140-6736(20)31100-4)

Tsang, Nicole Ngai Yung, Hau Chi So, Ka Yan Ng, Benjamin J. Cowling, Gabriel M. Leung, and Dennis Kai Ming Ip. 2021. “Diagnostic Performance of Different Sampling Approaches for SARS-CoV-2 RT-PCR Testing: A Systematic Review and Meta-Analysis.” *The Lancet Infectious Diseases* 0 (0). <https://doi.org/10.1016/S1473-3099(21)00146-8>.

Venkatachalam, Anuja. 2021. “3 Reasons Why India Could Be Underreporting COVID-19 Deaths.” Health Analytics Asia. May 6, 2021. [http://www.ha-asia.com/3-reasons-why-india-](http://www.ha-asia.com/3-reasons-why-india-could-be-underreporting-covid-19-deaths/) [could-be-underreporting-covid-19-deaths/](http://www.ha-asia.com/3-reasons-why-india-could-be-underreporting-covid-19-deaths/).

Verity, Robert, Lucy C Okell, Ilaria Dorigatti, Peter Winskill, Charles Whittaker, Natsuko Imai, Gina Cuomo-Dannenburg, et al. 2020. “Estimates of the Severity of Coronavirus Disease 2019: A Model-Based Analysis.” *The Lancet Infectious Diseases* 20 (6): 669–77. [https://doi.org/10.1016/S1473-3099(20)30243-7.](https://doi.org/10.1016/S1473-3099(20)30243-7)

Walker, Patrick G. T., Charles Whittaker, Oliver J. Watson, Marc Baguelin, Peter Winskill, Arran Hamlet, Bimandra A. Djafaara, et al. 2020. “The Impact of COVID-19 and Strategies for Mitigation and Suppression in Low- and Middle-Income Countries.” *Science* 369 (6502): 413–22. [https://doi.org/10.1126/science.abc0035.](https://doi.org/10.1126/science.abc0035)

Wang, Rui, Yuta Hozumi, Changchuan Yin, and Guo-Wei Wei. 2020. “Mutations on COVID-19 Diagnostic Targets.” *Genomics* 112 (6): 5204–13. [https://doi.org/10.1016/j.ygeno.2020.09.028.](https://doi.org/10.1016/j.ygeno.2020.09.028)

Ward, Helen, Graham S. Cooke, Christina Atchison, Matthew Whitaker, Joshua Elliott, Maya Moshe, Jonathan C. Brown, et al. 2021. “Prevalence of Antibody Positivity to SARS-CoV-2 Following the First Peak of Infection in England: Serial Cross-Sectional Studies of 365,000 Adults.” *The Lancet Regional Health – Europe* 4 (May). [https://doi.org/10.1016/j.lanepe.2021.100098.](https://doi.org/10.1016/j.lanepe.2021.100098)

Ward, Helen, Christina Atchison, Matthew Whitaker, Kylie EC Ainslie, Joshua Elliott, Lucy Okell, Rozlyn Redd, et al. 2020. “Antibody Prevalence for SARS-CoV-2 Following the Peak of the Pandemic in England: REACT2 Study in 100,000 Adults.” *Mimeo-MedRxiv*, August, 2020.08.12.20173690. <https://doi.org/10.1101/2020.08.12.20173690>.

World Health Organization (WHO). 2020a. “Naming the Coronavirus Disease (COVID-19) and the Virus That Causes It.” n.d. Accessed July 1, 2021. [https://www.who.int/emergencies/diseases/novel-coronavirus-2019/technical-](https://www.who.int/emergencies/diseases/novel-coronavirus-2019/technical-guidance/naming-the-coronavirus-disease-(covid-2019)-and-the-virus-that-causes-it) [guidance/naming-the-coronavirus-disease-(covid-2019)-and-the-virus-that-causes-it](https://www.who.int/emergencies/diseases/novel-coronavirus-2019/technical-guidance/naming-the-coronavirus-disease-(covid-2019)-and-the-virus-that-causes-it).

World Health Organization (WHO). 2020b. “Laboratory testing strategy recommendations for COVID-19: interim guidance, 21 March 2020.” World Health Organization. <https://apps.who.int/iris/handle/10665/331509>. License: CC BY-NC-SA 3.0 IGO

World Health Organization (WHO). 2020c. “Tracking SARS-CoV-2 Variants.” n.d. Accessed June 8, 2021. [https://www.who.int/activities/tracking-SARS-CoV-2-variants.](https://www.who.int/activities/tracking-SARS-CoV-2-variants)

World Health Organization (WHO). 2020d. “Medical Certification, ICD Mortality Coding, and Reporting Mortality Associated with COVID-19.” n.d. Accessed July 3, 2021. [https://www.who.int/publications-detail-redirect/WHO-2019-nCoV-mortality-reporting-](https://www.who.int/publications-detail-redirect/WHO-2019-nCoV-mortality-reporting-2020-1) [2020-1.](https://www.who.int/publications-detail-redirect/WHO-2019-nCoV-mortality-reporting-2020-1)

Xiao, Ai Tang, Yi Xin Tong, Chun Gao, Li Zhu, Yu Jie Zhang, and Sheng Zhang. 2020. “Dynamic Profile of RT-PCR Findings from 301 COVID-19 Patients in Wuhan, China: A Descriptive Study.” *Journal of Clinical Virology: The Official Publication of the Pan American Society for Clinical Virology* 127 (June): 104346. <https://doi.org/10.1016/j.jcv.2020.104346>.

Yadav, Radha, Arup Acharjee, Akanksha Salkar, Renuka Bankar, Viswanthram Palanivel, Sachee Agrawal, Jayanthi Shastri, Sanjeev V. Sabnis, and Sanjeeva Srivastava. 2021. “Mumbai Mayhem of COVID-19 Pandemic Reveals Important Factors That Influence Susceptibility to Infection.” *EClinicalMedicine* 35 (May). <https://doi.org/10.1016/j.eclinm.2021.100841>.

Yang, Wan, and Jeffrey Shaman. 2021. “Epidemiological Characteristics of Three SARS- CoV-2 Variants of Concern and Implications for Future COVID-19 Pandemic Outcomes.” *Mimeo-MedRxiv*, May, 2021.05.19.21257476. <https://doi.org/10.1101/2021.05.19.21257476>.

Yousaf, Anna R, Lindsey M Duca, Victoria Chu, Hannah E Reses, Mark Fajans, Elizabeth M Rabold, Rebecca L Laws, et al. 2020. “A Prospective Cohort Study in Nonhospitalized Household Contacts With Severe Acute Respiratory Syndrome Coronavirus 2 Infection: Symptom Profiles and Symptom Change Over Time.” *Clinical Infectious Diseases*, no. ciaa1072 (July). <https://doi.org/10.1093/cid/ciaa1072>.

Zhang, Lizhou, Cody B. Jackson, Huihui Mou, Amrita Ojha, Haiyong Peng, Brian D. Quinlan, Erumbi S. Rangarajan, et al. 2020. “SARS-CoV-2 Spike-Protein D614G Mutation Increases Virion Spike Density and Infectivity.” *Nature Communications* 11 (1): 6013. [https://doi.org/10.1038/s41467-020-19808-4.](https://doi.org/10.1038/s41467-020-19808-4)

Zimmer, Carl. 2021. “How the ‘Alpha’ Coronavirus Variant Became So Powerful.” *The New York Times*, June 7, 2021, sec. Health. [https://www.nytimes.com/2021/06/07/health/covid-](https://www.nytimes.com/2021/06/07/health/covid-alpha-uk-variant.html) [alpha-uk-variant.html.](https://www.nytimes.com/2021/06/07/health/covid-alpha-uk-variant.html)
